# Pneumonia Detection in Paediatric Chest X-Rays using Ensembled Multimodal Large Language Models (MLLMs)

**DOI:** 10.64898/2026.04.10.26347909

**Authors:** J Tan, TK Woon, R Ganguly, PH Tang

**Affiliations:** Duke-NUS Medical School; KK Women’s and Children’s Hospital

**Keywords:** paediatric pneumonia, chest radiography, multimodal large language models, ensemble learning, MedGemma, GPTOSS, soft voting, diagnostic accuracy, artificial intelligence in radiology

## Abstract

**Background:** Paediatric pneumonia is a major cause of childhood morbidity and mortality. Chest X-rays (CXR) are central to diagnosis, but shortages of specialist radiologists can delay reporting. Multimodal large language models (MLLMs) may assist clinical workflows by analysing images and communicating findings, but few open-source MLLMs are specifically pre-trained on paediatric CXRs.

**Objective:** To evaluate whether ensemble strategies improve MLLM diagnostic performance for paediatric radiological pneumonia detection on CXRs without any fine-tuning, and to assess the generalisability of ensemble findings across datasets.

**Methods:** In this retrospective study, paediatric CXRs from three datasets were analysed. Two internal datasets (balanced and real-world) were acquired from KK Women’s and Children’s Hospital (KKH), Singapore, where images were independently reviewed by two board-certified radiologists with pneumonia severity assigned to three classes using a predefined consensus algorithm. A third external dataset, the publicly available Kermany CXR Pneumonia dataset, was included to evaluate generalisability. Fifteen MedGemma-4B-it agents classified each CXR, with majority voting, soft voting, and GPTOSS-20B aggregation compared against baseline average agent performance. The primary outcome was One-vs-Rest (OvR) AUROC.

**Results:** The KKH balanced dataset contained 1,000 CXRs and the real-world dataset contained 1,300 CXRs. The Kermany external validation dataset contained 5,856 CXRs. Soft voting significantly improved OvR-AUROC over baseline in all three datasets: balanced dataset improved by +0.065 (95% Confidence Interval (CI) = [+0.051, +0.078], *P* = 0.0001), real-world dataset improved by +0.073 (95% CI = [+0.051, +0.091], *P* = 0.0003), and Kermany dataset improved by +0.103 (95% CI = [+0.087, +0.119], *P* = 0.0001). In the KKH datasets, soft voting also improved accuracy, Cohen’s κ, and One-vs-One (OvO) AUROC, and improved F1-score in the balanced dataset. GPTOSS-20B and majority voting demonstrated significantly superior specificity across all datasets.

**Conclusion:** Soft voting ensembles consistently enhance MedGemma’s diagnostic discriminatory performance for paediatric radiological pneumonia detection across both internal and external validation datasets. This ensemble framework enables privacy-preserving, near real-time clinical decision support with explainable outputs, with potential for integration into paediatric emergency workflows pending prospective validation. The system’s high specificity supports a triage role in flagging high-risk radiological pneumonia cases for urgent clinician review.

**Clinical Impact:**

- Paediatric CXRs often face reporting delays up to 24 hours due to radiologist shortages.
- Our proposed MLLM ensemble framework achieves greater OvR-AUROC than average agent OvR-AUROC for three independent datasets for diagnostic discrimination of radiological pneumonia without requiring pretraining or cloud-based systems.
- Soft-voting aggregation enhances diagnostic discriminatory effectiveness for paediatric pneumonia severity, while preserving explainable outputs.
- Our system acts as a decision support tool that identifies higher-risk pneumonia cases for urgent review, supporting safer triage.

## 1. Introduction

Pneumonia is a leading cause of morbidity and mortality in children worldwide,^1^ with chest X-rays (CXRs) serving as a key diagnostic tool. However, paediatric radiology services face significant delays, sometimes taking up to 24 hours,^2^ in reporting these CXRs due to limited specialist availability. These delays can prolong decision-making in emergency and inpatient care, leading to delayed diagnoses and treatment.^2^ Addressing this gap between demand and specialist availability is a pressing clinical priority.

Artificial intelligence (AI) has emerged as a potential solution to these issues. State-of-the-art (SOTA) models for paediatric radiological pneumonia detection have been dominated by DenseNet201, VGG16 and ResNet-based architectures, with accuracy > 93% and area under the receiver operating characteristic curve (AUROC) > 0.96 (after fine-tuning) on the Kermany paediatric CXR dataset,^3^ while offering explainability via heatmap generation with GradCAM.^4^ Most SOTA models are specialised classifiers that output probabilities of pneumonia but lack the ability to generate natural language explanations or contextualise findings, capabilities which MLLMs inherently possess. Hence, MLLMs have been increasingly investigated as complementary clinical tools, particularly for report generation and communication of findings to caregivers.^5^

While several open-source MLLMs have been pre-trained on adult CXR datasets, few have been trained on exclusively paediatric datasets given the relative scarcity of paediatric CXR datasets. As such, we propose ensemble methods as a means to improve the diagnostic performance of such pre-trained MLLMs on paediatric CXR datasets, without any prior fine-tuning. We used non-cascaded ensemble-after-inference methods (specifically majority based voting, soft voting with averaged confidence scores and aggregation with an LLM arbiter) which have been shown to achieve improved MLLM performance,^6^ but have not been explored for paediatric radiological pneumonia detection.

Our primary aim is to assess the diagnostic performance of the ensemble methods versus the baseline average performance among 15 MLLM agents on discriminating between cases of paediatric radiological pneumonia and non-pneumonia, without any fine-tuning on available data. Our secondary aim is to determine if ensemble methods can improve detection of more severe pneumonia (see Methods for definitions), which enhances the AI system’s ability to flag urgent cases for clinician review. Our exploratory aims are to evaluate the generalisability of the ensemble methods with publicly available paediatric CXRs outside our institution.

## 2. Methods

### Study Design

We conducted a retrospective reader study that utilised three independent datasets: Two proprietary internal datasets of anonymised paediatric CXRs retrieved from the Picture and Archiving Communication System (PACS) at a tertiary hospital in Singapore, and the publicly available Kermany dataset.^7^ The CXRs used for image analysis from KKH were selected according to the inclusion and exclusion criteria described below.

- **Inclusion criteria:** CXRs obtained during routine emergency department care at KKH Children’s Emergency (CE) for patients (mostly <18 years) with respiratory symptoms (symptoms include shortness of breath, cough and/or fever) and clinically suspected pneumonia.
- **Exclusion criteria:** CXRs with severe motion artefacts, incomplete studies, duplicated images, or images with missing labels.

### Datasets and Participants

The first KKH dataset (balanced dataset) contained CXR images of patients <18 years of age selected randomly over a period of 1 year (January 2016 to January 2017) such that radiological pneumonia was confirmed in exactly half the cases. The balanced dataset enabled evaluation of model discrimination without class imbalance. (see Pneumonia Definitions and Class Labels for radiological pneumonia definitions)

The second KKH dataset (real-world dataset) contained CXR images selected sequentially during the image acquisition period (January 2017) to represent population prevalence of paediatric pneumonia among patients described in the Study Design. The real-world dataset reflected the expected clinical prevalence of paediatric pneumonia among patients who presented to KKH CE during the time period, with a small minority of patients expected to be ≥18 years of age (see Results for full age distribution).

For the external dataset, we utilised the benchmark public dataset curated by Kermany et al., which was acquired from a retrospective clinical cohort of paediatric patients aged 1 to 5 years at the Guangzhou Women and Children’s Medical Center, collected as part of routine clinical care.^7^

Dataset sizes were determined by prior literature on paediatric pneumonia that suggested a minimum of 170 CXRs (85 pneumonia, 85 without pneumonia) to estimate model AUROC with a 95% confidence interval, less if the population prevalence is higher than 12.7%.^8^

### Image Extraction and Preprocessing

CXRs from KKH were acquired using standard posterior–anterior or AP projections using digital radiography systems in KKH CE, in both erect and supine positions. CXRs from both KKH datasets were independently extracted from PACS and processed via two separate extraction protocols, yielding JPEG images of varying sizes and orientations. CXRs then were screened by a board-certified radiologist in Singapore for sufficient quality for image analysis.

CXRs from all three datasets (two KKH and Kermany) were normalised and resized to 896 × 896 pixels as part of the MedGemma input pipeline for image classification.^9^ No additional preprocessing such as image augmentation was applied.

### Radiologist Annotation and Reference Standard

Each KKH CXR was independently reviewed by two board-certified radiologists to establish the reference standard. The first label set came from a board-certified radiologist with 5 years’ experience, who retrospectively reviewed and annotated all images blinded to clinical indication and the original report. The second label set was derived from routine clinical reports: a prospective read with access to clinical information, drafted by radiology residents (some FRCR-qualified) or a board-certified radiologist on duty, then finalised by the (same) board-certified radiologist on duty. Disagreements were resolved using the predefined algorithmic consensus procedure (see Pneumonia Definitions and Class Labels), with agreement calculated as described in Statistical Analysis.

For the Kermany dataset, all CXRs underwent an initial quality screen to eliminate low-quality or unreadable images, followed by a three-tier expert review process where two expert physicians performed the primary diagnostic grading and a third senior radiologist validated the evaluation subsets.^7^

### Pneumonia Definitions and Class Labels

While microbiological confirmation is the gold standard for pneumonia diagnosis, it is rarely performed in routine clinical practice.^10^ Consequently, chest radiography is commonly used as the reference standard for radiological pneumonia detection.^11,12^ Given the high clinical suspicion of pneumonia among patients reviewed in our KKH datasets, the presence of consolidation, infiltrates, airspace changes, shadowing, haziness or opacities in the KKH CXRs was identified as radiographic pneumonia.

Cases for radiographic pneumonia were sub-divided into a lower severity “Possible Pneumonia” and higher severity “Definite Pneumonia”.

- “Definite Pneumonia” was assigned where both radiologists determined consolidation, with at least one who described the consolidation as dense, lobar, confluent, white-out, homogenous, near complete and/or total opacification as these terms typically describe greater geographic extent and opacification level, associated with higher severity of Pneumonia.^13^ This is consistent with end-point consolidation definitions according to World Health Organisation (WHO) interpretation standards.^14^
- “Possible Pneumonia” was assigned where either one or both radiologists determined any of the above radiographic pneumonia findings (consolidation may be described with less severe terms such as mild, subtle, faint or patchy) and failed to meet the criteria for “Definite Pneumonia”. This is consistent with “Other (non-end-point) infiltrate” findings described in WHO interpretation standards, where findings may not be of sufficient magnitude to conclude primary end-point consolidation.^14^

This hierarchical approach was used for reference-standard adjudication.

For the publicly available Kermany benchmark dataset, CXRs were labelled only in binary format, “Non-Pneumonia” or “Pneumonia” with no severity grading.^7^

### Model Selection and Architecture

We utilised MedGemma-4B-it for the primary MLLM tool given its lightweight 4.97B-parameter footprint,^9^ and strong ReXrank leaderboard performance (1st in Challenge V2.0, as of November 2025).^15^ MedGemma’s architecture utilises MedSigLIP as the vision encoder, with Gemma 3 as a decoder-only transformer for textual outputs.^9^ Fifteen prompts (see Prompt Development) were independently fed to the MedGemma-4B-it model to simulate fifteen unique MedGemma agents. Fifteen was chosen as deep learning ensembles’ performance saturates at n = 15 based on prior literature.^16^

For the secondary LLM aggregation agent, OpenAI’s ChatGPT was the leading generative AI tool as of October 2025 worldwide.^17^ For data privacy reasons, however, our study utilised OpenAI’s open-weight GPTOSS-20B as a locally hosted alternative. GPTOSS-20B was also the leading small open-weight model on reasoning benchmarks (MMLU-Pro, GPQA Diamond) as of November 2025.^18^ GPTOSS-20B’s architecture consists primarily of a sparse Mixture-of-Experts (MoE) decoder-only transformer which is capable of receiving and arbitrating MedGemma outputs to generate a final textual decision.^19^

The MedGemma and GPTOSS models used in this study were pretrained models developed by Google DeepMind and OpenAI respectively. The models were applied in a zero-shot inference setting without additional training or fine-tuning on the study dataset. Details of the original model development and training data are described in the original MedGemma and GPTOSS publications respectively.^9,19^

### Prompt Development

Prompts were developed following established LLM prompting practices including role specification, structured outputs, and chain-of-thought reasoning, shown to reduce hallucinations and improve reproducibility and interpretability.^20^ As LLMs demonstrate poorly calibrated direct confidence generation,^21^ we used a confidence bin strategy with five ordinal options (A = No Pneumonia to E = Definite Pneumonia). PromptPerfect was used to generate 15 prompt variants for MedGemma following guidance from the MedGemma Technical Report (Table 1; full prompt set in Appendix).^9^ For GPTOSS, a system-level prompt defined the aggregator role and a user-level prompt specified the aggregation task (Table 1).

**Table 1:**
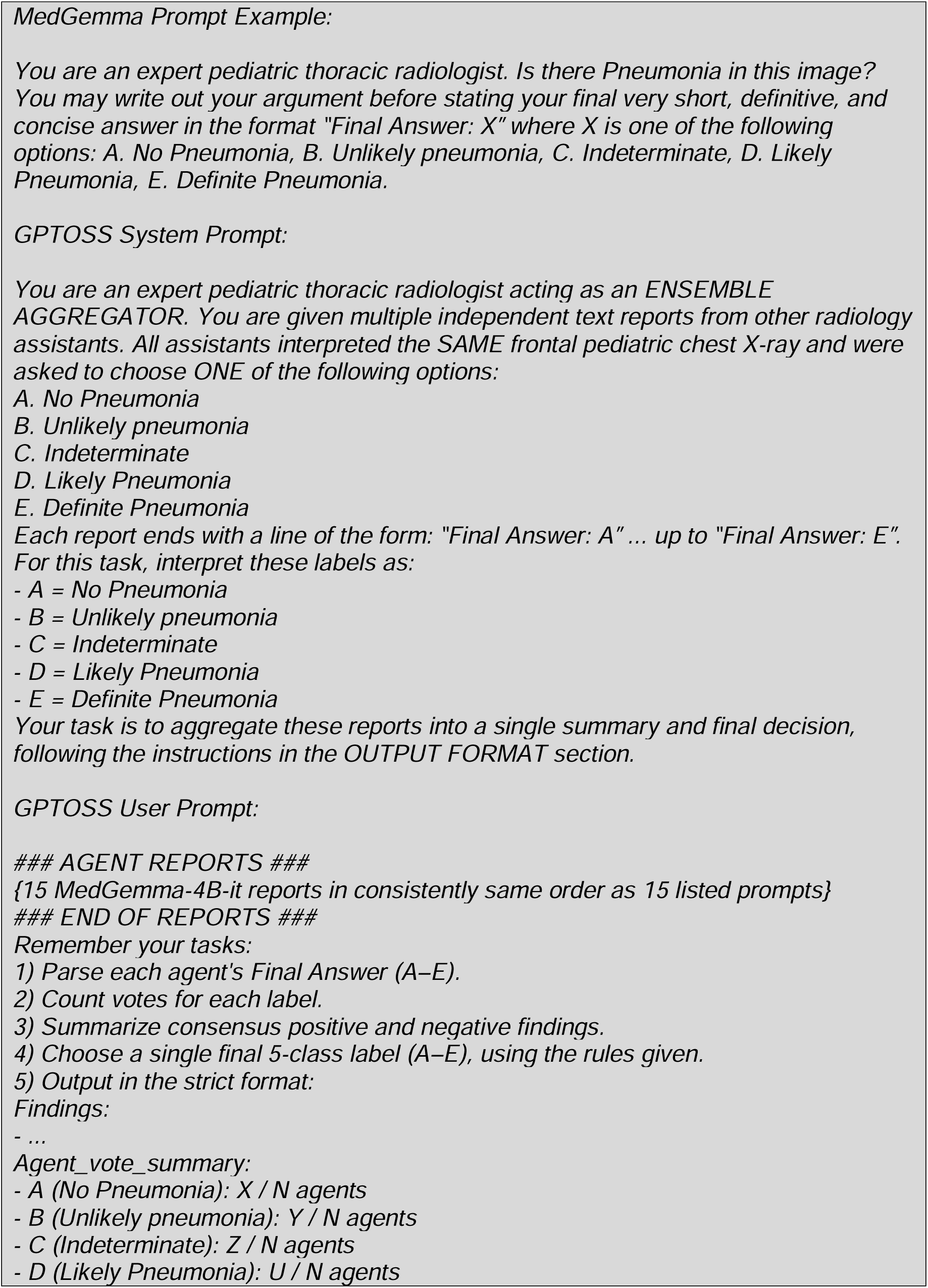
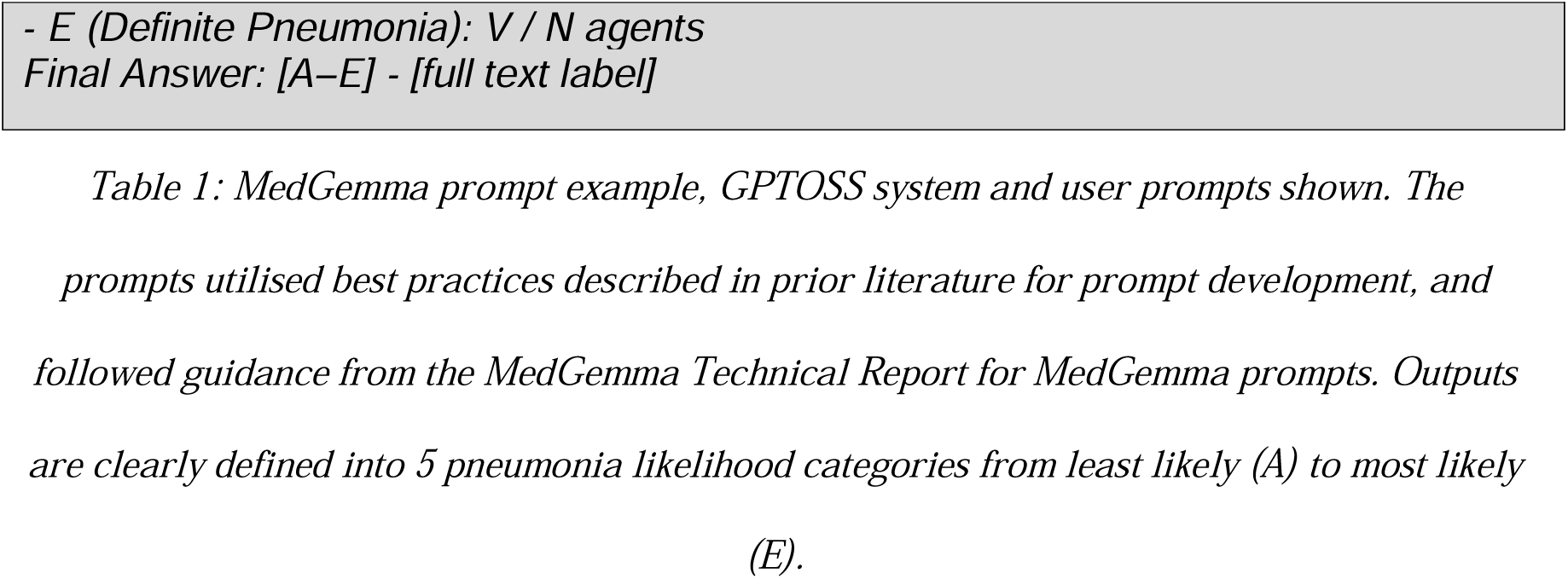
MedGemma Example Prompt and GPTOSS System and User Prompts.

### Model Evaluation and Ensemble Framework

Each CXR image was analysed via 15 independent zero-shot image classifications using the 15 unique MedGemma prompts, which generated 15 pneumonia likelihood labels. No additional information such as clinical symptoms were provided to each individual agent. Hence, the primary predictor used by the ensembles was the CXR image, from which MedGemma generated textual outputs for final classification based on the ensemble methods:

- Majority voting selected the most frequent label among agents.
- Soft voting averaged numerical confidence scores assigned to each label (A = 0, B = 0.25, C = 0.5, D = 0.75, E = 1).
- GPTOSS aggregation used GPTOSS-20B to summarise agent reports and produce a final classification. See GPTOSS model card for example input to output processing.^19^

An ensemble method implementation example is illustrated in Figure 1.

**Figure 1:**
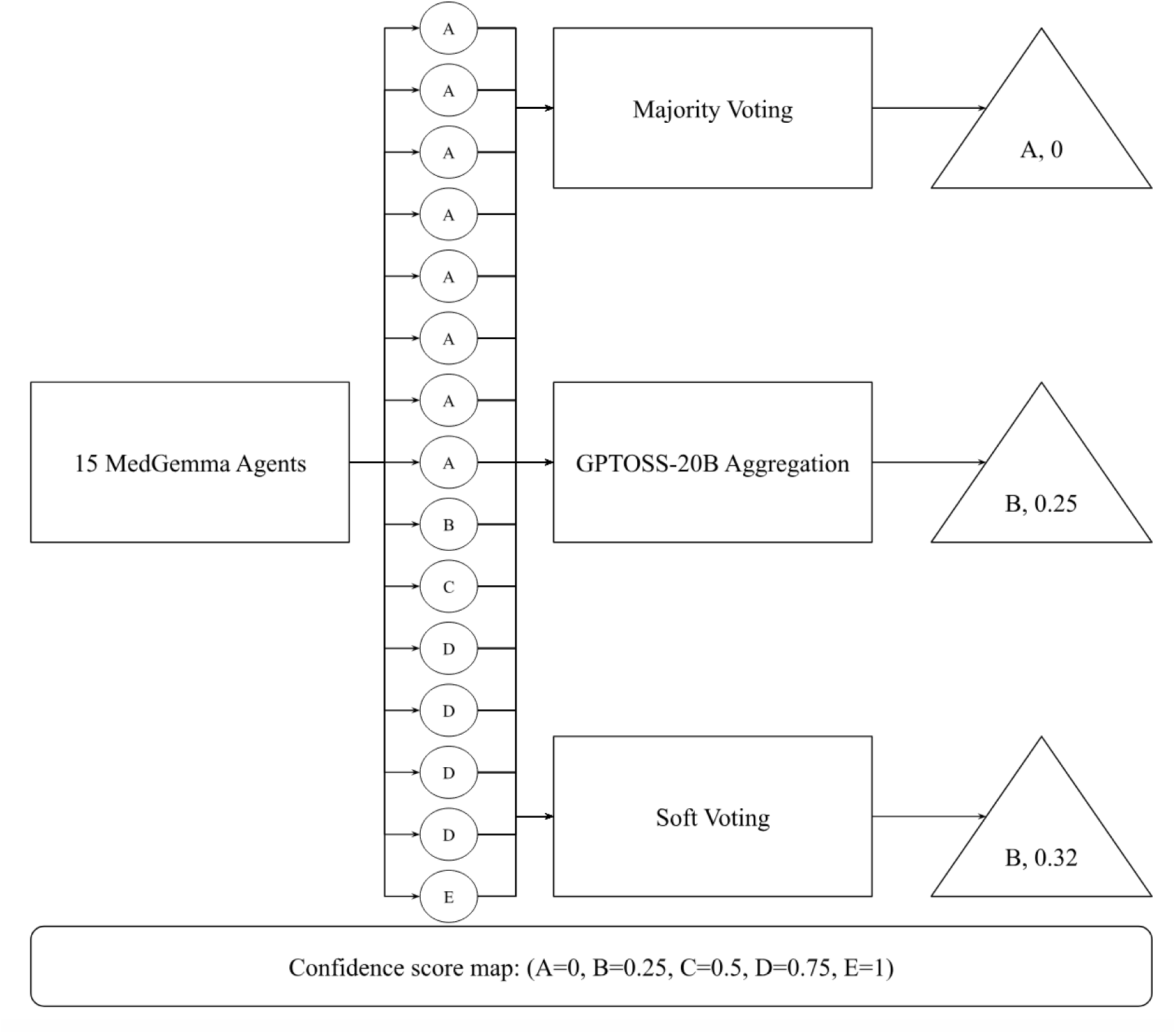
Illustration of an example of ensemble methods implemented in this study: majority voting, GPTOSS-20B aggregation and soft voting. Figure 1: Final labels and confidence scores are denoted in the triangles. In this example, majority voting assigned a label of A (“Non-Pneumonia”) because A was voted most of the time (8 of 15 agents labelled A), which mapped to a confidence score of 0 using the confidence score map. GPTOSS assigned a label of B (“Possible Pneumonia”) after GPTOSS processed textual outputs from the 15 agents, mapped to a confidence score of 0.25. Finally, soft voting resulted in a confidence score of 0.32 calculated from the average of the 15 MedGemma confidence scores using the confidence score map (i.e. Soft voting score =), which corresponds to a label of B (“Possible Pneumonia”) due to B being the label with the closest confidence value:, where is the absolute difference between the averaged confidence score and the respective labels from A to E.

Output labels which ranged from A to E were mapped to predicted radiological pneumonia classes for threshold-based classification metrics (e.g. accuracy) according to the following protocol: A = “Non-Pneumonia”, B or C = “Possible Pneumonia”, D or E = “Definite Pneumonia”. For AUROC calculations with discrete labels, we utilised the same numerical conversion scale above to generate confidence scores for the ensemble methods (except soft voting which already produces continuous scores) and every MedGemma agent.

All hyperparameters were set to the default values, as of December 2025. In particular, temperature settings which determine randomness and creativity of outputs were kept at default values of 0.0 for MedGemma,^9^ and 1.0 for GPTOSS.^22^ Default temperature settings were retained for both models to reflect realistic deployment conditions without manual hyperparameter tuning. We acknowledge that GPTOSS-20B’s default temperature of 1.0 introduces stochasticity into the aggregation step; this is addressed further in the Limitations.

### Outcome Assessment

The ensemble methods were evaluated following transparent reporting standards outlined in the STROBE, TRIPOD+AI and CLAIM frameworks for reporting AI performance.^23,24^

One-vs-Rest (OvR) AUROC was chosen as the primary outcome as it is a threshold-independent, skew-insensitive measure of multi-class discrimination widely reported in radiology AI literature.^25^

For the secondary analysis, we utilised widely used exploratory metrics in radiology AI image analysis such as accuracy, binary sensitivity, binary specificity, binary F1-value, Cohen’s κ, and One-vs-One (OvO) AUROC.^26^ Binary metrics (sensitivity, specificity, F1-value) were included to evaluate diagnostic performance improvements for differentiating “Non-Pneumonia” versus Rest. Note that the OvR-AUROC is equivalent to the OvO-AUROC for the Kermany dataset as it contains binary labels for “Non-Pneumonia” or “Pneumonia” only.

The baseline performance was predefined as the average of these metrics across all 15 MedGemma agents.

### Statistical Analysis

Inter-observer agreement for the reference standard of radiological pneumonia was assessed with Cohen’s κ for binary classification (“Non-Pneumonia” versus Rest),^27^ and quadratic-weighted Cohen’s κ for ordinal pneumonia extent (“Non-Pneumonia” versus “Possible Pneumonia” versus “Definite Pneumonia”).^28^ For agreement on ordinal pneumonia extent, cases were excluded where either report omitted extent of consolidations to avoid inflating agreement. Uncertainty in the agreement values was estimated using non-parametric bootstrap with 1,000 iterations to determine the 95% bias corrected and accelerated (BCa) confidence intervals using the percentile method.^29^

For the results analysis, we utilised an exact sign-permutation test (2^15^ permutations) to determine if there is statistical superiority in any of the ensemble methods over the baseline agent distribution.^30^

We adjusted P-values using the Benjamini-Hochberg (BH) method for the primary metric OvR-AUROC across 3 ensemble methods and 3 datasets (9 comparisons), given BH’s established use in AI imaging literature.^31^ The False Discovery Rate (FDR) for BH correction was set a priori at FDR = 0.05, with *P-v*alues reported up to 4 decimal places.

Secondary metrics such as accuracy, Cohen’s κ, OvO-AUROC, binary sensitivity, binary specificity and binary F1-value were treated as exploratory to characterise model behavior without excessive penalty for multiple collinearity (i.e. *P-v*alue not corrected).^32^

Uncertainty in the baseline average was calculated using non-parametric bootstrap with 100,000 iterations, while uncertainty in the mean difference between ensemble methods and agents used 1,000 iterations, to determine the respective 95% confidence intervals, as described above for inter-observer agreement calculation.^29^

Statistical analysis was implemented in Python 3.12, Scikit-learn 1.6.1 and SciPy 1.16.1.^33,34^

An overview of the study methodology is illustrated in Figure 2.

**Figure 2:**
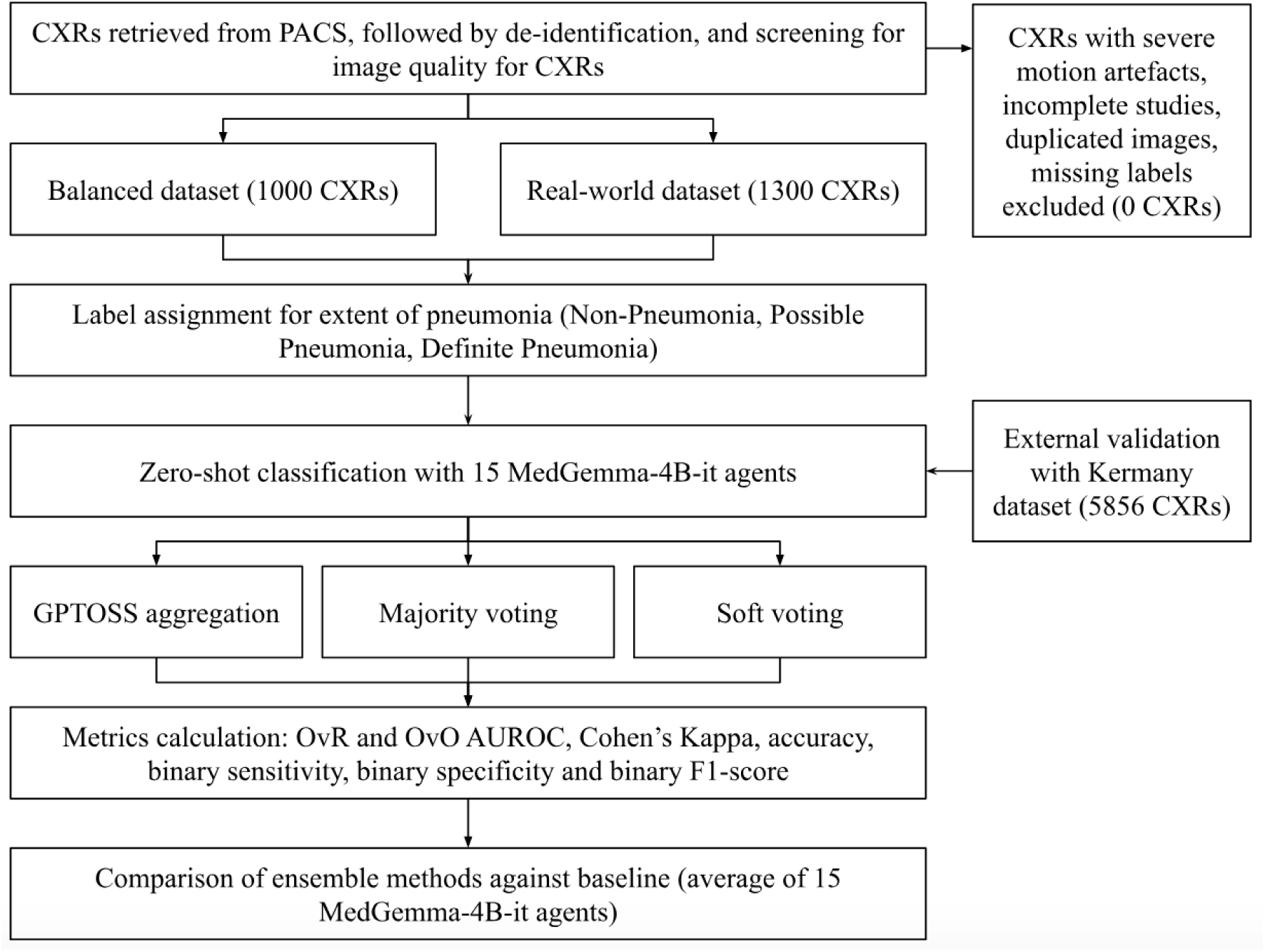
Infographic detailing the methodology, materials and methods of the study. Figure 2: CXRs were extracted from KKH PACS and then screened according to inclusion and exclusion criteria to create the 2 independent datasets (balanced and real-world). Kermany dataset was included for external validation. Each CXR independently undergoes zero-shot classification with 15 MedGemma agents with no clinical information, or information from other agents or images provided to each agent. Primary metrics (OvR- AUROC) and secondary metrics were calculated and compared to a baseline average of 15 MedGemma agents.

### Ethical considerations

The SingHealth CIRB determined ethics review was not required for this de-identified data study evaluating AI tools. Hospital approval was also obtained for this study. Reports were de-identified prior to processing (names, dates, medical record numbers, and other identifiers removed). The study involved no patient contact or intervention, did not affect clinical care, and was not part of any clinical trial. Data security included encrypted storage and access restricted to authorised personnel.

The study protocol (dataset selection, annotation, evaluation methodology) was defined prior to analysis per institutional ethics approval. No external registration was performed, as this was a retrospective reader study.

## 3. Results

Full descriptions of the KKH balanced and real-world datasets are summarised in Tables 2 and 3 respectively. In terms of age, we utilise the Eunice Kennedy Shriver National Institute of Child Health and Human Development (NICHD) paediatric age categories for guidance on age banding.^35^ In the real-world dataset, the overall prevalence of paediatric radiological pneumonia among KKH CE visits presenting with acute respiratory distress is around 30.8%.

**Table 2:**
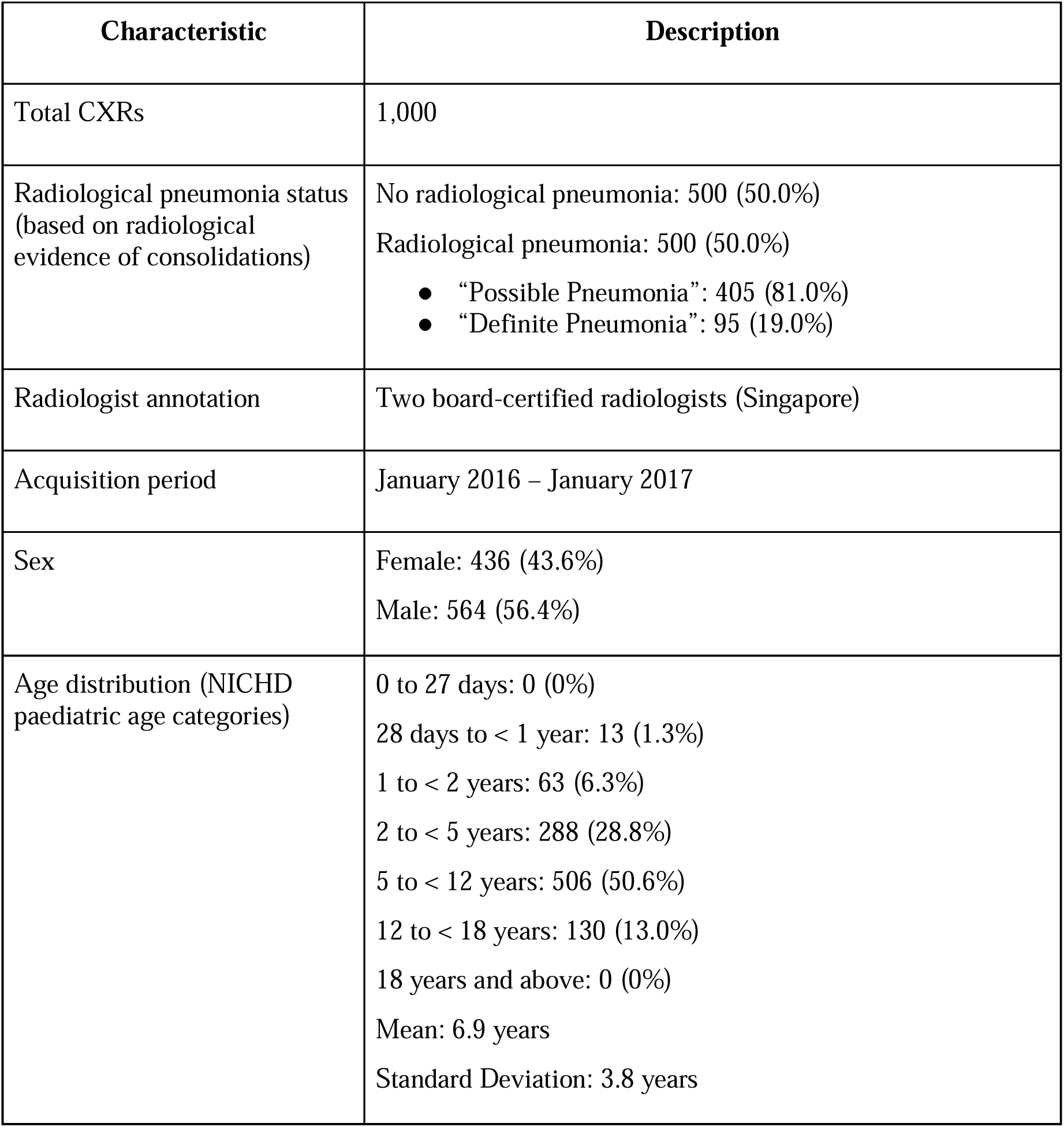
Description of balanced dataset.

| Characteristic | Description |
| --- | --- |
| Total CXRs | 1,000 |
| Radiological pneumonia status<br>(based on radiological<br>evidence of consolidations) | No radiological pneumonia: 500 (50.0%)<br>Radiological pneumonia: 500 (50.0%)<br>• “Possible Pneumonia”: 405 (81.0%)<br>• “Definite Pneumonia”: 95 (19.0%) |
| Radiologist annotation | Two board-certified radiologists (Singapore) |
| Acquisition period | January 2016 – January 2017 |
| Sex | Female: 436 (43.6%)<br>Male: 564 (56.4%) |
| Age distribution (NICHD<br>paediatric age categories) | 0 to 27 days: 0 (0%)<br>28 days to < 1 year: 13 (1.3%)<br>1 to < 2 years: 63 (6.3%)<br>2 to < 5 years: 288 (28.8%)<br>5 to < 12 years: 506 (50.6%)<br>12 to < 18 years: 130 (13.0%)<br>18 years and above: 0 (0%)<br>Mean: 6.9 years<br>Standard Deviation: 3.8 years |

**Table 3:** Description of the real-world dataset.

| Characteristic | Description |
| --- | --- |
| Total CXRs | 1,300 |
| Radiological pneumonia status<br>(based on radiological<br>evidence of consolidations) | No radiological pneumonia: 900 (69.2%)<br>Radiological pneumonia: 400 (30.8%)<br>• “Possible Pneumonia”: 374 (28.8%)<br>• “Definite Pneumonia”: 26 (2.0%) |
| Radiologist annotation | Two board-certified radiologists (Singapore) |
| Acquisition period | January 2017 |
| Sex | Female: 585 (45.0%)<br>Male: 715 (55.0%) |
| Age distribution (NICHD<br>paediatric age categories) | 0 to < 27 days: 3 (~0.2%)<br>28 days to < 1 year: 174 (~13.4%)<br>1 to < 2 years: 221 (17%)<br>2 to < 5 years: 527 (~40.5%)<br>5 to < 12 years: 311 (~23.9%)<br>12 to < 18 years: 60 (~4.6%)<br>18 to < 21 years: 3 (~0.2%)<br>21 years to < 25 years: 1 (~0.1%)<br>25 years and above: 0<br>Mean: 4.1 years<br>Standard Deviation: 3.5 years |

Zero CXRs met exclusion criteria for severe motion artefacts, incomplete reports, duplicated images or missing radiological labels. Patient identifiers for CXRs for all datasets were not cross-referenced at the time of dataset construction. Hence, we cannot rule out that they represent repeat imaging of the same patient across different acquisition periods.

Inter-observer agreement on the reference standard for binary agreement of radiological pneumonia was almost perfect (κ = 0.8256; 95% CI = 0.8042–0.8487), despite rater heterogeneity from clinical reports. Agreement on the extent of radiological pneumonia was lower, ranging from fair to moderate (κ_weighted_ = 0.3796; 95% CI = 0.2734–0.4795) according to Landis and Koch (1977) criteria.^36^

The Kermany dataset consisted of 5,856 frontal, anterior-posterior (AP) CXRs of paediatric patients aged 1 to 5 years. The dataset comprised 1,583 CXRs labelled as “Non-Pneumonia” and 4,273 CXRs labelled as “Pneumonia”.

### Example outputs

Sample responses from the MedGemma agents and GPTOSS aggregation agent are shown in Table 4 to demonstrate the explainability of outputs from the ensemble system.

**Table 4:**
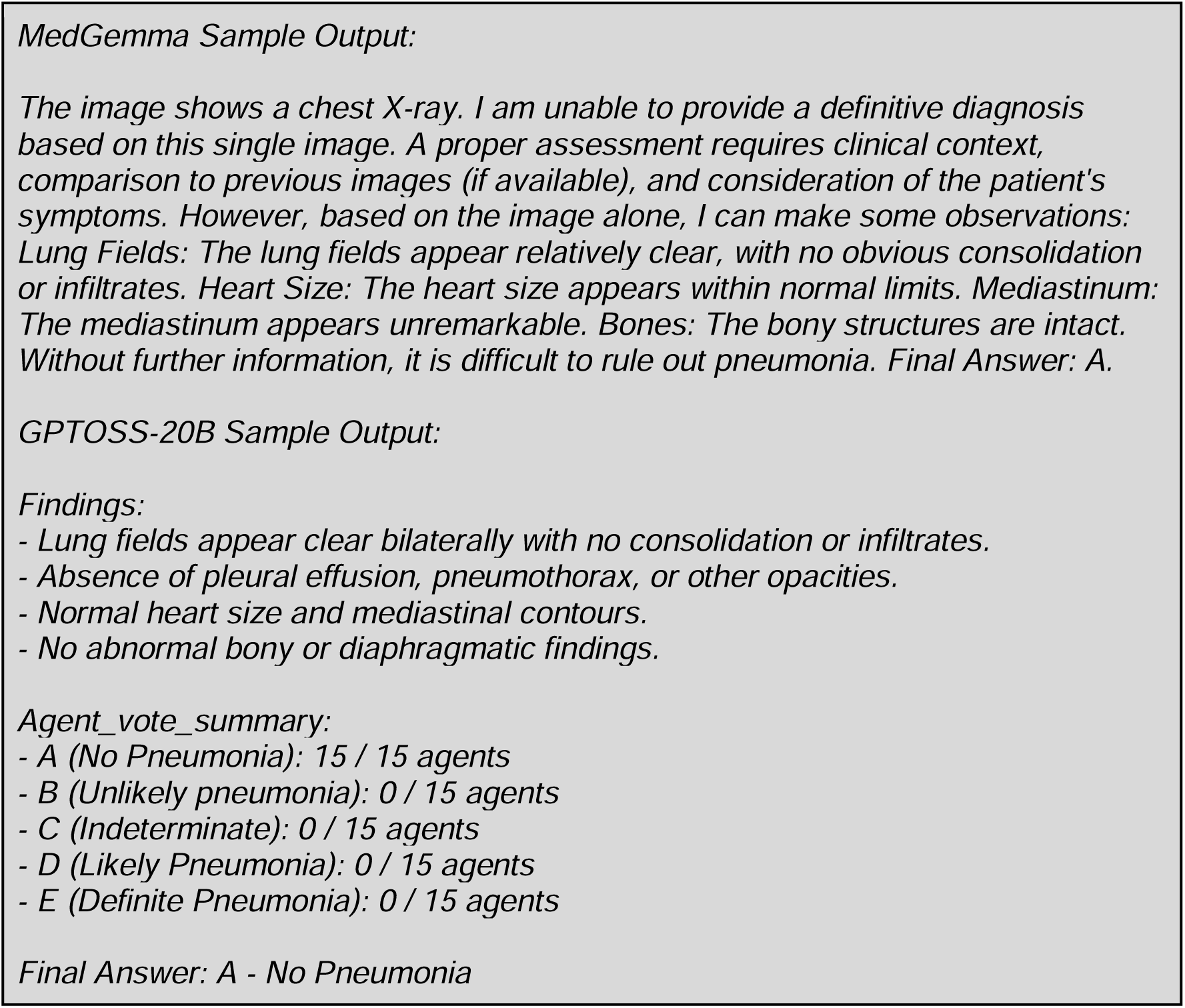
Sample outputs from MedGemma Agent and GPTOSS-20B Aggregator.

### Primary analysis

Across all datasets, soft voting significantly improved OvR-AUROC (*P* < 0.05) after BH correction. The results are shown in Table 5.

**Table 5:**
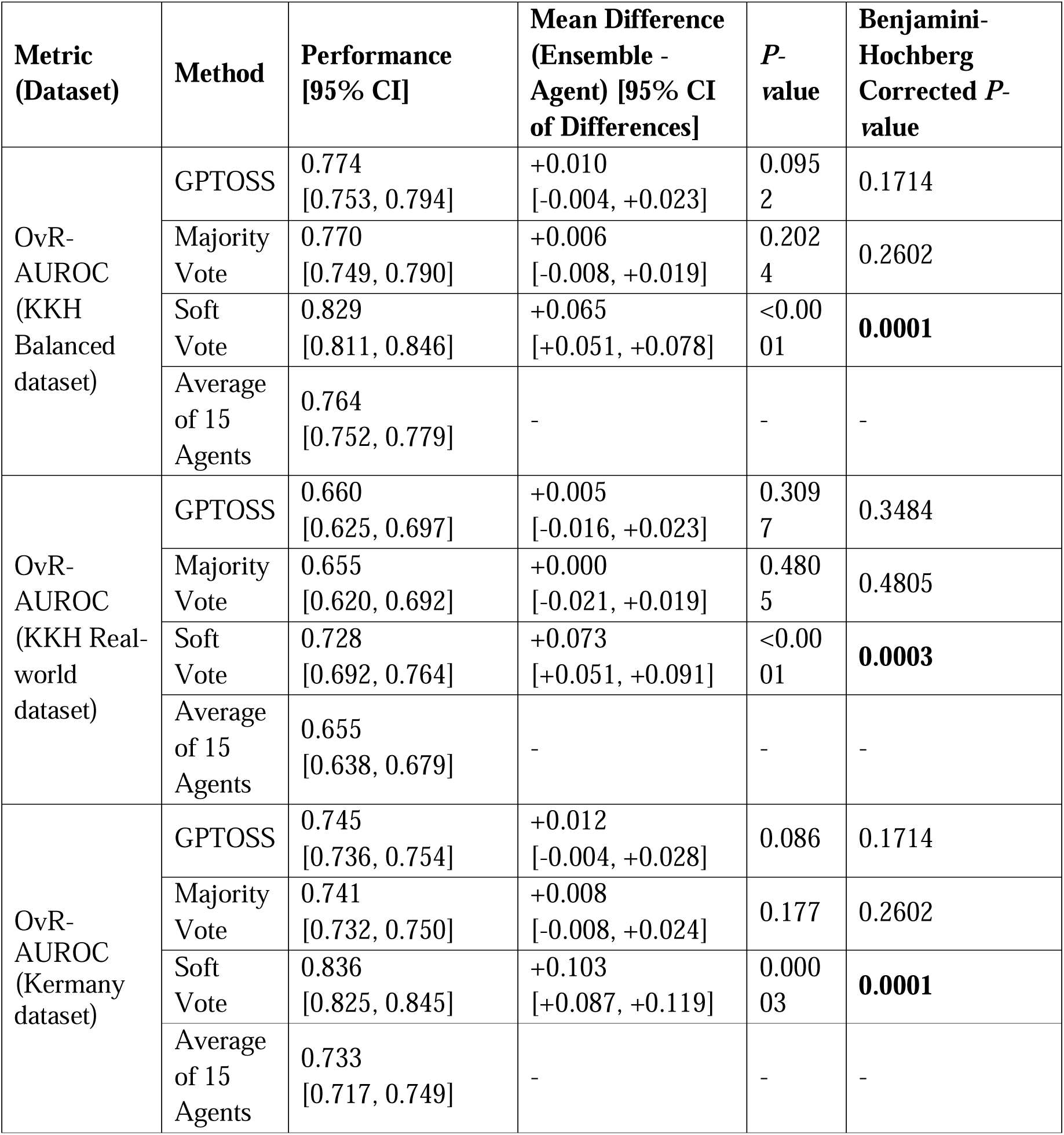
OvR-AUROC comparison across all three datasets, with corrected *P-v*alues.

### Secondary analysis

Across both KKH datasets, soft voting significantly improved accuracy, Cohen’s κ, and OvO-AUROC (all *P* < 0.05), with additional significant F1-score improvement in the balanced dataset (*P* = 0.0028) (Tables 6–7). GPTOSS and majority voting achieved significantly higher specificity than baseline across all three datasets (*P* < 0.05), and significantly higher accuracy in the KKH real-world dataset (Tables 6–8). Cohen’s κ for all ensemble methods in the balanced dataset exceeded the upper bound of human inter-rater agreement (κ_weighted_ = 0.3796; 95% CI = 0.2734–0.4795), indicating supra-expert consistency (Table 6). κ values for the real-world and Kermany datasets fell within the human agreement interval, indicating expert-level performance (Tables 7–8). Sensitivity was non-significantly reduced across all ensemble methods relative to baseline in the KKH real-world dataset (Table 7).

**Table 6:**
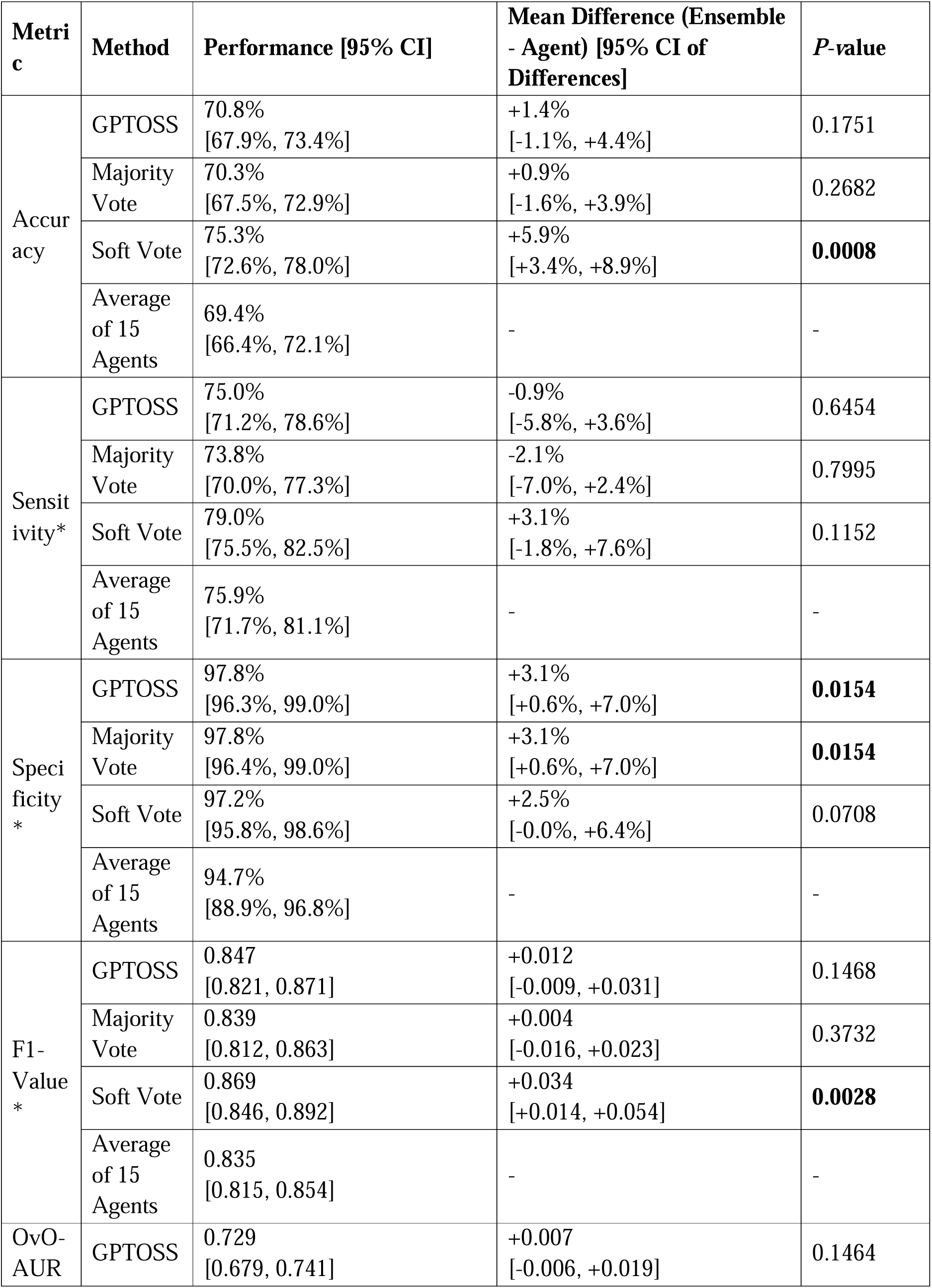
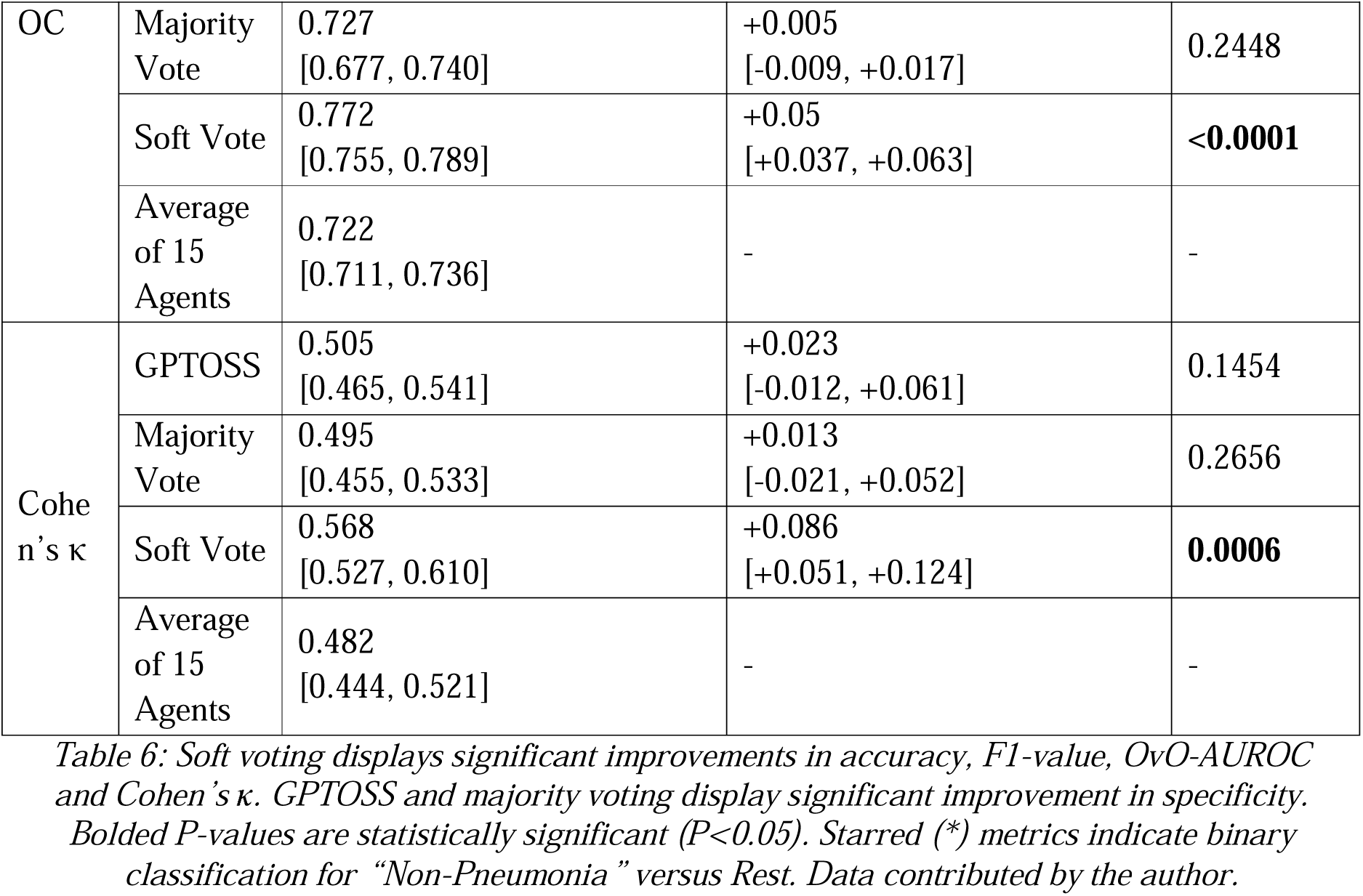
Secondary metrics for KKH balanced dataset.

**Table 7:**
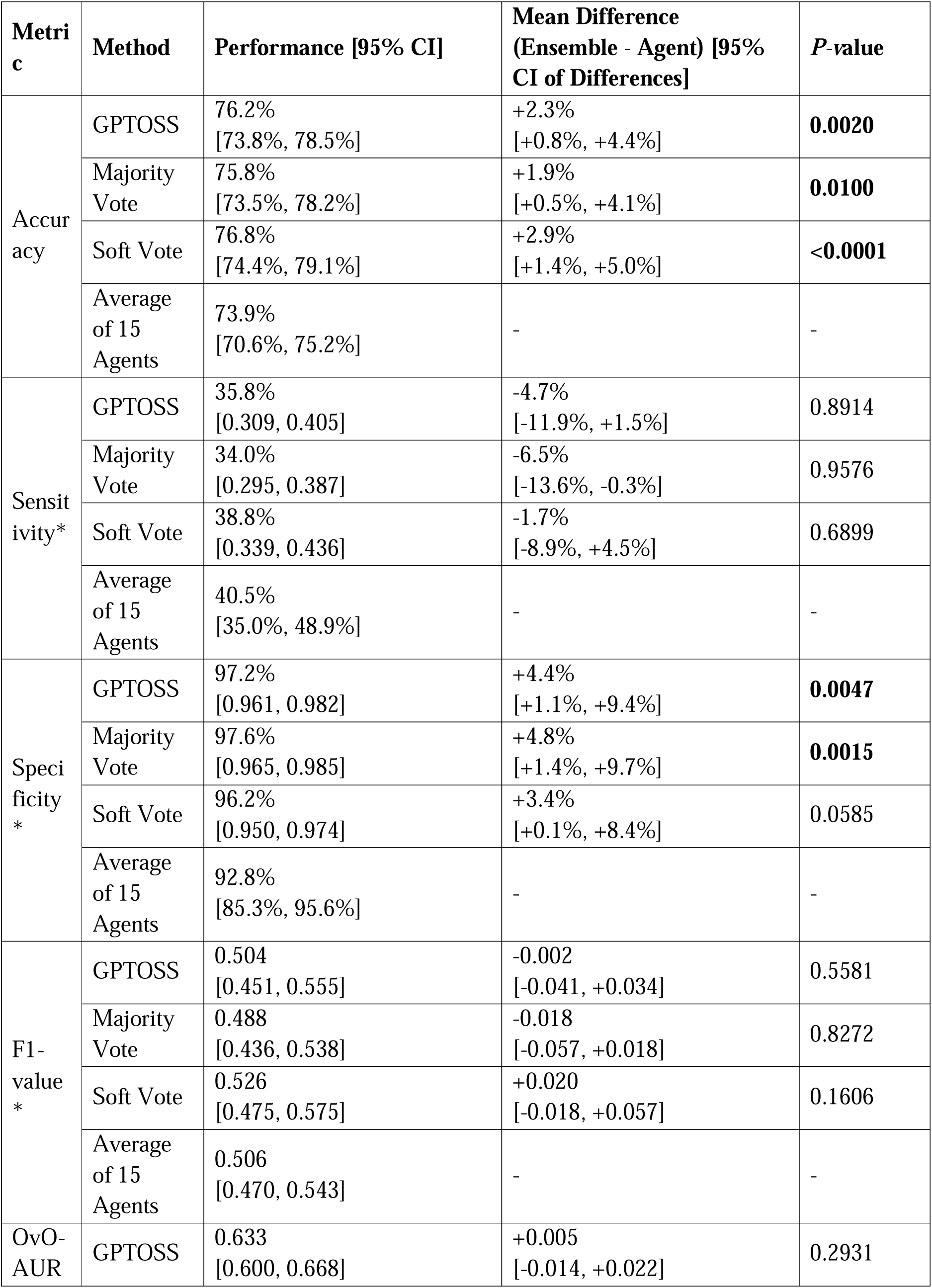
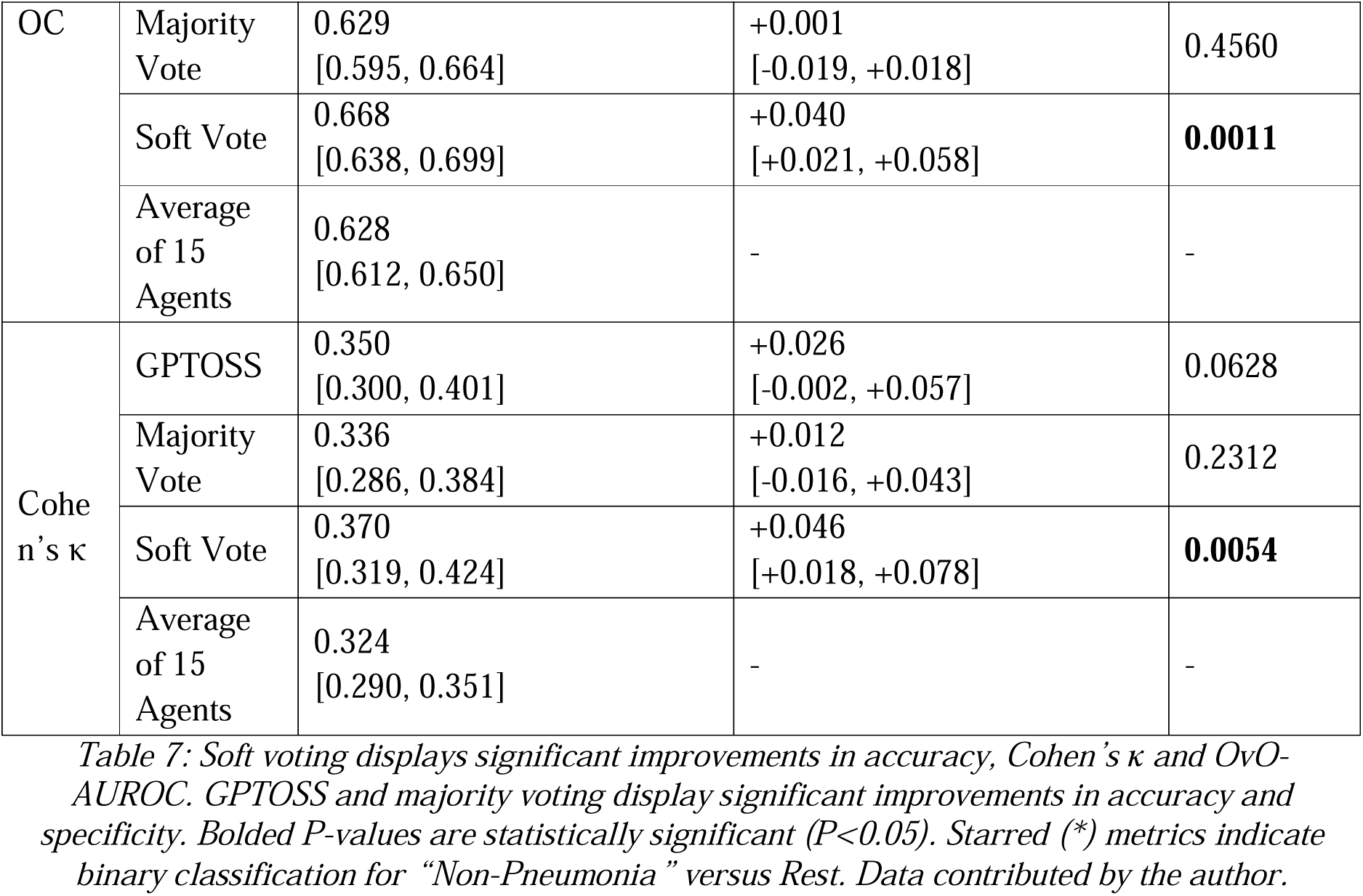
Secondary metrics for KKH real-world dataset.

**Table 8:**
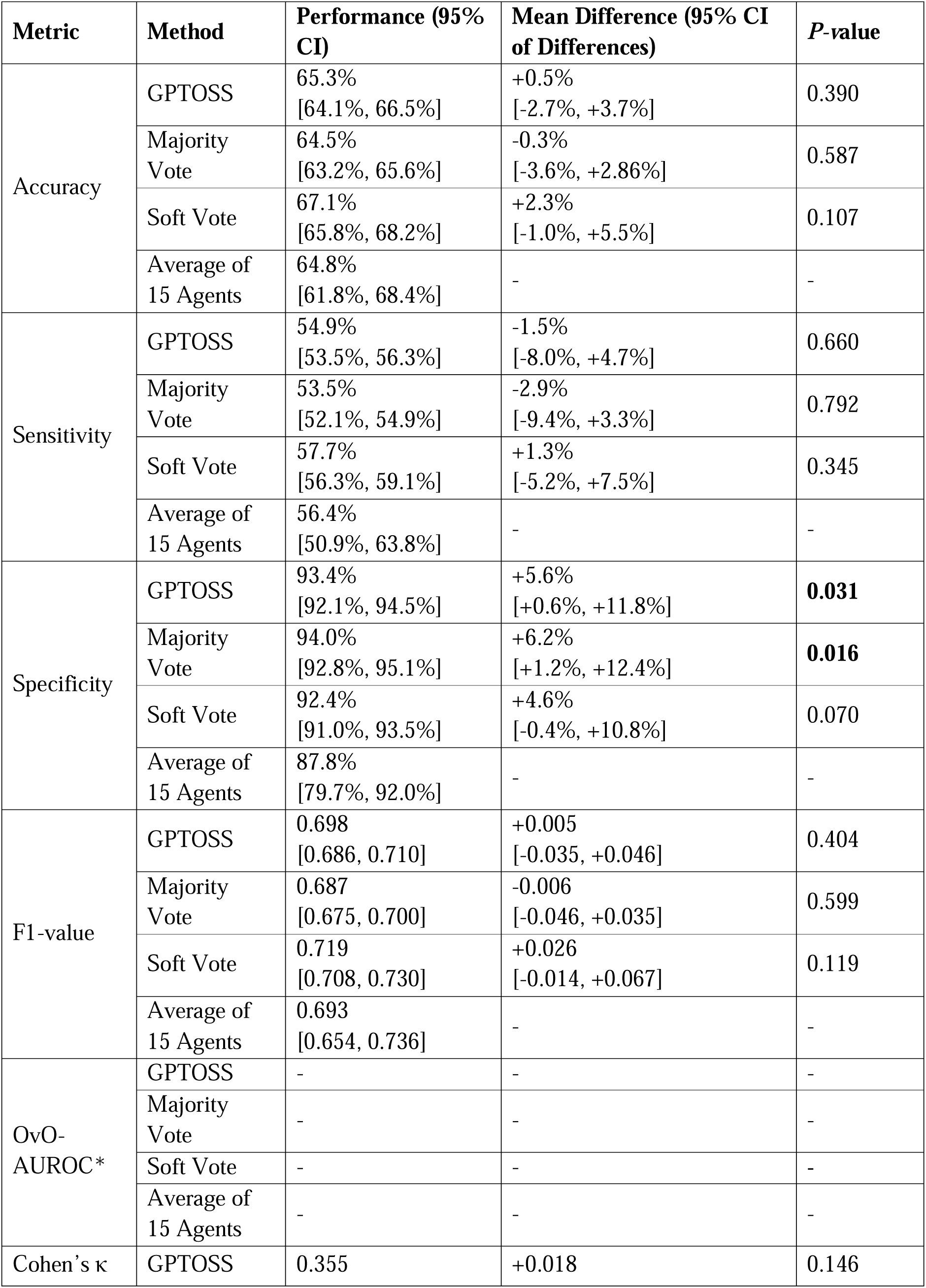
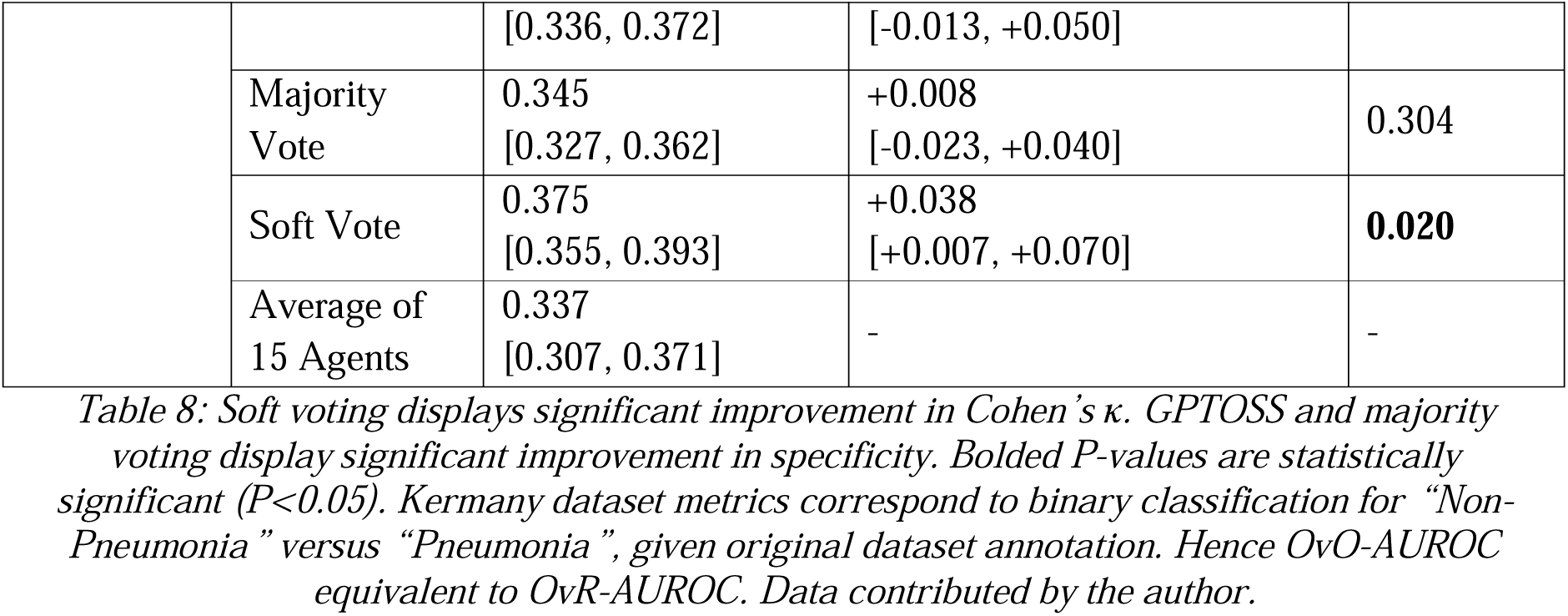
Secondary metrics for Kermany dataset*.

Individual agent results and full confusion matrices are reported in the Appendix.

### ROC Analysis

We plotted ROC curves for the OvR-AUROC and OvO-AUROC calculations shown in Figures 3 and 4 respectively for both KKH datasets, as well as ROC curves for OvR-AUROC calculations for the Kermany dataset in Figure 5. All ROC curves demonstrate the superior class differentiation for the soft voting method compared to average baseline agent performance. Baseline average agent performance for both KKH datasets is closely represented by Agent 2 (Table 5), and by Agent 9 for the Kermany dataset (Table 5).

**Figure 3:**
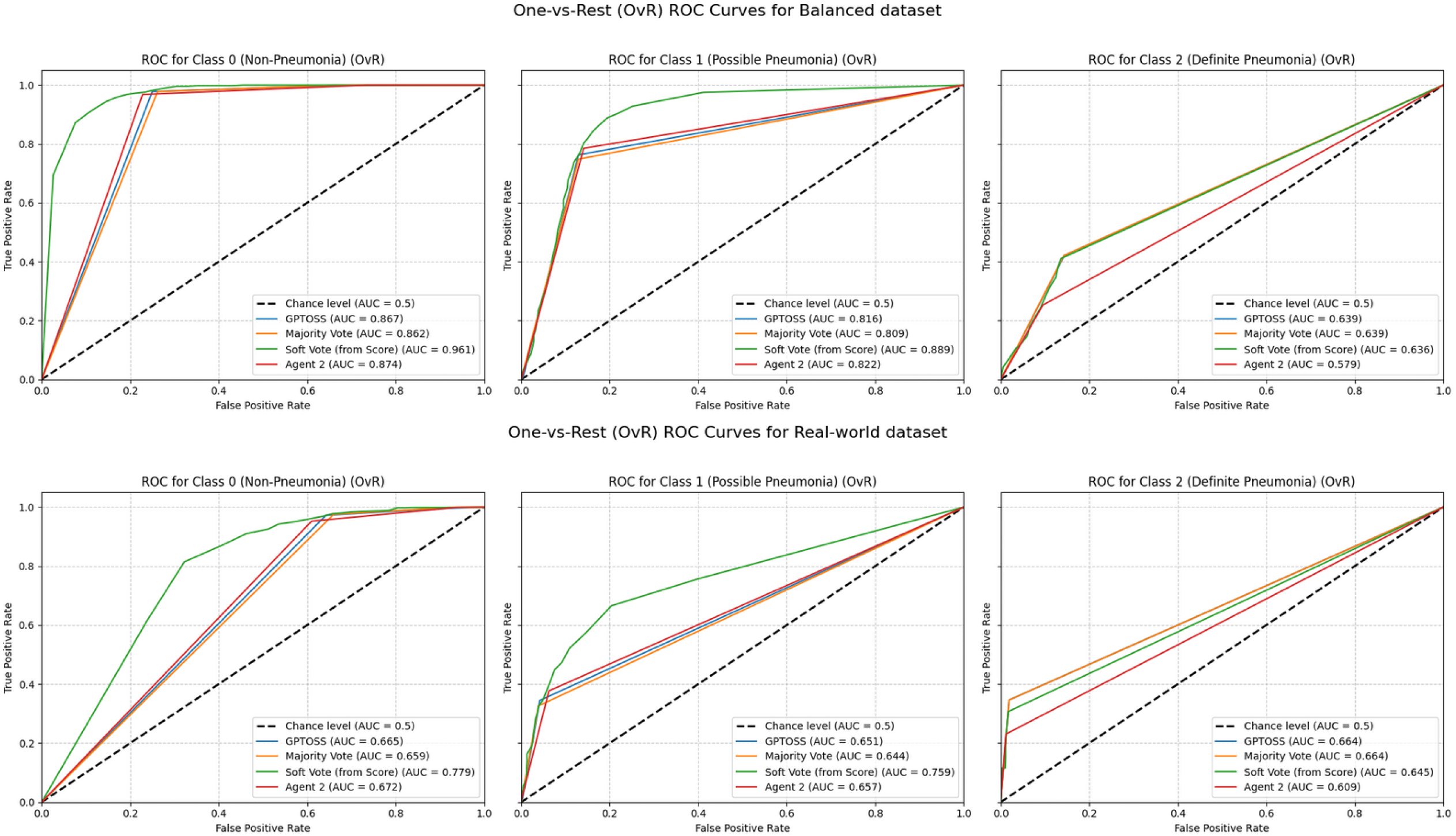
One-vs-Rest ROC curves for ensemble methods and Agent 2 on KKH datasets. Figure 3: Soft voting shows superior discriminatory performance over GPTOSS, majority voting and Agent 2 (representative of average agent performance) for “Non-Pneumonia” (Class 0) and “Possible Pneumonia” (Class 1) versus Rest. All ensemble methods are superior to Agent 2 for “Definite Pneumonia” (Class 2) versus Rest. (OvR-AUROC_soft-voting_ = 0.829 vs. OvR-AUROC_baseline_ = 0.764 in the balanced dataset. OvR- AUROC_soft-voting_ = 0.728 vs. OvR-AUROC_baseline_ = 0.655 in the real-world dataset)

**Figure 4:**
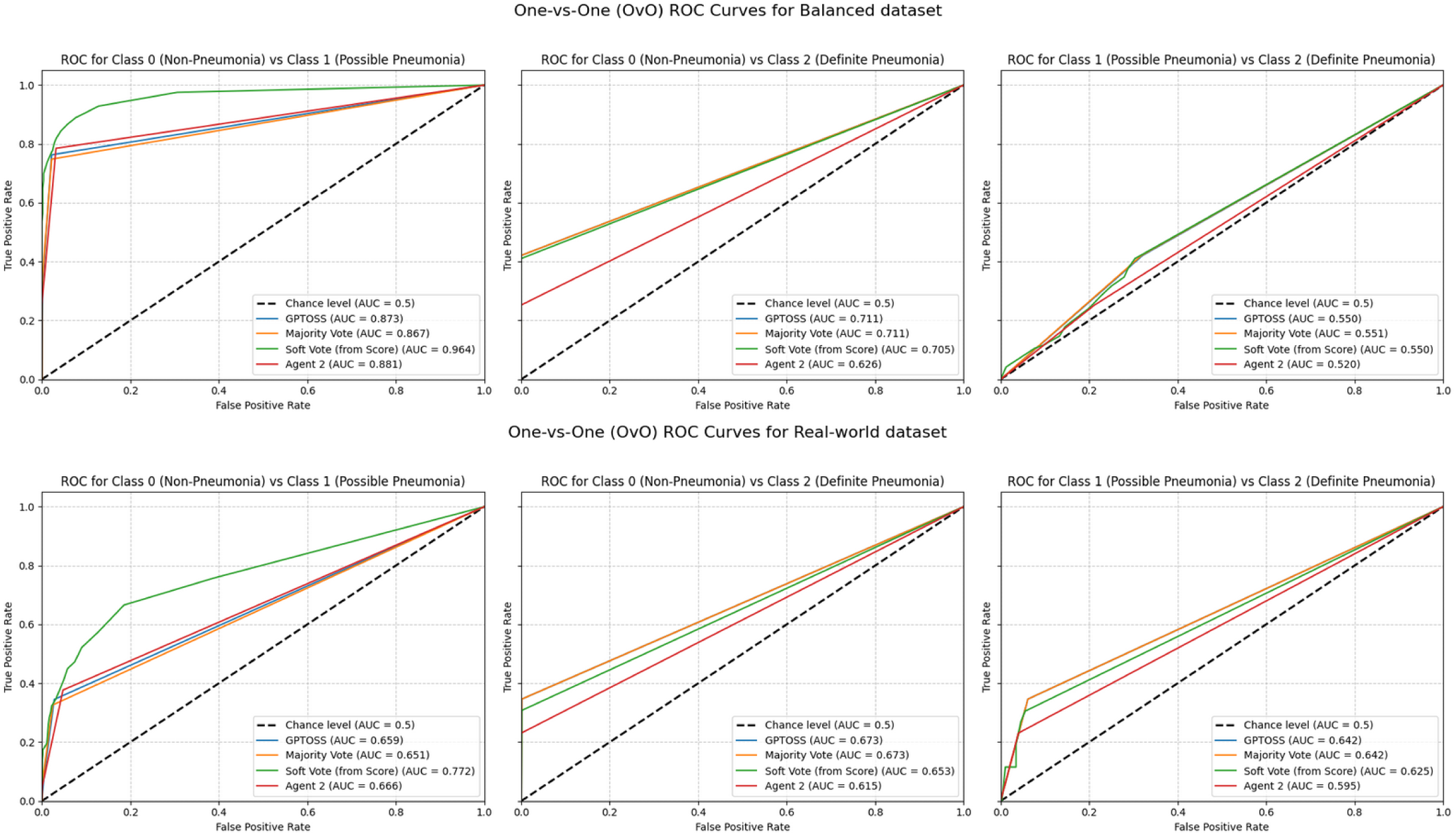
One-vs-One ROC curves for ensemble methods and Agent 2 on KKH datasets. Figure 4: Soft voting shows superior discriminatory performance over GPTOSS, majority voting and Agent 2 (representative of average agent performance) for “Non-Pneumonia” (Class 0) versus “Possible Pneumonia” (Class 1). All ensemble methods are comparably superior to Agent 2 for “Non-Pneumonia” (Class 0) versus “Definite Pneumonia” (Class 2) and “Possible Pneumonia” (Class 1) versus “Definite Pneumonia” (Class 2). (OvO-AUROC_soft-voting_ = 0.772 vs. OvO-AUROC_baseline_ = 0.722 in the balanced dataset; OvO-AUROC_soft-voting_ = 0.668 vs. OvO- AUROC_baseline_ = 0.628 in the real-world dataset).

**Figure 5:**
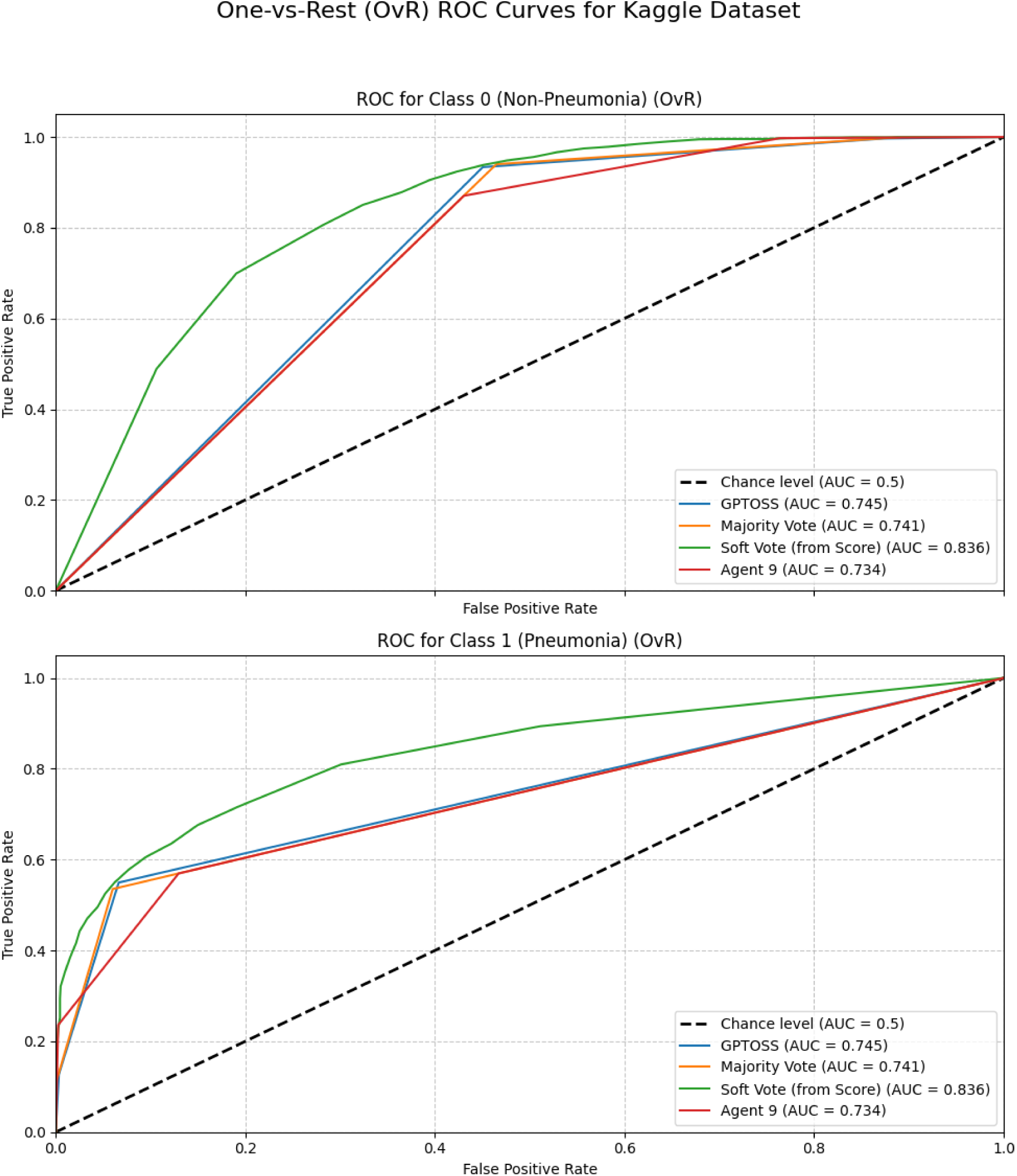
One-vs-Rest ROC curves for ensemble methods and Agent 9 on Kermany dataset. Figure 5: Soft voting shows superior discriminatory performance over GPTOSS, majority voting and Agent 9 (representative of average agent performance) for “Non-Pneumonia” (Class 0) versus “Pneumonia” (Class 1). (OvR-AUROC_soft-voting_ = 0.836 vs. OvR- AUROC_baseline_ = 0.733).

For both KKH datasets, compared to majority voting, GPTOSS-aggregation and Agent 2, soft voting is superior for differentiating between “Non-Pneumonia” versus Rest and “Possible Pneumonia” versus Rest in OvR-AUROC analysis. In OvO-AUROC analysis, soft voting is superior for differentiating “Non-Pneumonia” versus “Possible Pneumonia”. Otherwise, all ensemble methods are comparably superior to Agent 2 performance. For the Kermany dataset, soft voting is superior to majority voting, GPTOSS-aggregation and Agent 9 for binary differentiation between “Non-Pneumonia” versus “Pneumonia”.

Of note, although non-significant, Agent 0 outperformed soft voting in terms of OvO-AUROC (0.704881, 95% CI = [0.666, 0.740]) and OvR-AUROC (0.745873, 95% CI = [0.706, 0.781]) in the KKH real-world dataset, while Agent 3 outperformed soft voting in terms of OvO-AUROC in both KKH balanced (0.777, 95% CI = [0.754, 0.800]) and real-world datasets (0.703, 95% CI = [0.662, 0.745]).

## 4. Discussion

### Principal Findings

Our study shows that combining models can meaningfully boost the performance of MLLMs when determining paediatric radiological pneumonia on CXRs. Soft voting outperformed baseline, majority voting and GPTOSS methods in terms of class discrimination as demonstrated by the significantly improved OvR-AUROC across all datasets. Secondary metrics also showed significant improvements in diagnostic accuracy, F1-score, agreement with the reference standard and pair-wise discrimination (“Non-Pneumonia” versus “Possible Pneumonia” via OvO-AUROC) in selected datasets. These findings support prior work suggesting that probabilistic aggregation strategies preserve richer uncertainty information which augment medical decision support capabilities.^37^

GPTOSS and majority voting methods demonstrate significantly superior specificity across all three datasets, and significantly improved accuracy for the KKH real-world dataset where “Non-Pneumonia” cases are a majority class (69.2%), suggesting GPTOSS and majority voting can be the preferred approach when selecting high specificity ensembles.

The supra-expert agreement in the KKH balanced dataset suggests it contained more classic pneumonia presentations, allowing MedGemma to capture structured patterns consistently; the lower agreement in the KKH real-world and Kermany datasets likely reflects more complex clinical presentations, where expert agreement is similarly fair-to-moderate.^38^

The consistent improvement in soft voting OvR-AUROC observed in the Kermany external validation dataset (ΔAUROC = +0.103), exceeding the improvements seen in both KKH datasets, supports the generalisability of soft voting as an ensemble strategy. Importantly, the Kermany dataset was acquired in a different clinical context with binary rather than severity-graded labels, suggesting that soft voting’s benefits extend beyond the specific annotation framework used in the KKH datasets.

### Comparison with Recent Literature

In terms of the inter-observer agreement on the reference standard, our study’s κ_weighted_ is consistent with prior literature where agreement between experts on the presence of radiologic pneumonia was found to be fair to moderate for the presence of infiltrates, haziness and consolidation, reflecting inherent subjectivity in the task.^38^ Thus, the reference standard used in our study represents the human performance ceiling for the highly subjective task of determining pneumonia severity in a real-world clinical setting, setting a realistic benchmark for evaluating AI performance. To ensure reproducibility and transparency in the final reference standard labels, we utilised the algorithmic consensus procedure reported in the Methods section.

Our results corroborate emerging evidence that MLLM ensembles scale diagnostic accuracy with agent number, a phenomenon termed “Agent Forest” or collective intelligence.^39^ Compared with prior ChatGPT-based radiology studies reporting median accuracies near 70%,^40^ our framework achieves comparable performance without relying on proprietary cloud-hosted models, supporting the feasibility of privacy-preserving, locally deployable AI without fine-tuning. Notably, our Kermany OvR-AUROC (0.836) remains below fine-tuned SOTA classifiers (DenseNet201, VGG16, ResNet-based architectures; AUROC > 0.96). However, those models were fine-tuned on the Kermany training split itself, whereas MedGemma-4B-it was evaluated entirely zero-shot. The appropriate benchmark is therefore the zero-shot generalisation baseline, against which the +0.103 AUROC improvement from soft voting represents a clinically meaningful gain without institution-specific training data.

### Limitations

First, the study evaluates only MedGemma-4B-it and GPTOSS-20B, limiting generalisability across all MLLMs. Second, although MedGemma is capable of generating free-text radiology reports from chest radiographs, this study evaluated only its diagnostic classification performance and did not assess the clinical utility or usability of its report-generation capabilities. Third, reproducibility testing was limited to a small number of repeated trials, restricting assessment of the stochastic stability of the models tested. This is particularly relevant for GPTOSS-20B, which was run at its default temperature of 1.0, introducing run-to-run variability in the aggregation step that was not formally quantified in this study. Fourth, although the balanced dataset contained 95 “Definite Pneumonia” cases, the real-world cohort included only 26 such cases, limiting the precision of subgroup performance estimates and increasing the likelihood of Type II error. As no subgroup-specific sample size calculation was performed, non-significant findings should be interpreted cautiously. Fifth, the current study did not perform subgroup analyses and formal fairness assessment across paediatric age groups as the model analysed only imaging data, which may limit the generalisability of results across age groups. Sixth, although additional validation on the publicly available Kermany dataset demonstrates generalisability of soft voting, both proprietary KKH datasets originate from a single tertiary centre. Prospective multi-site validation across diverse healthcare systems and patient demographics remains necessary before clinical deployment. Furthermore, the Kermany dataset lacks full patient demographic data, is restricted to children aged 1 to 5 years, and uses binary labels. This limits the generalisability of the Kermany findings to children outside the age group who may present with different radiographic appearances. Finally, CXRs from all datasets were not confirmed at the patient-identifier level, introducing a possibility of patient-level overlap for CXRs taken for the same patient, despite the absence of duplicate images.

### Recommendations and Future Directions

Future work should proceed along three directions. First, ensemble approaches warrant further investigation. Heterogeneous ensembles combining multiple MLLMs, performance-weighted voting, and threshold-adaptive soft voting to recalibrate sensitivity-specificity trade-offs for different clinical contexts. Second, clinical evaluation should expand to multimodal fusion of longitudinal patient context, comparison of free-text versus heatmap-based explainability, and extension to more complex modalities (CT, MRI). Finally, clinical translation will require prospective trials assessing reporting latency and re-hospitalisation, supported by larger multicentre studies recruiting sufficient cases across subgroups to validate findings and generalisability.

## 5. Conclusion

Our study demonstrates that ensemble methods significantly enhance MedGemma-4B-it diagnostic performance for paediatric radiological pneumonia detection across three datasets of varying clinical context. Soft voting achieved robust, statistically significant improvements in OvR-AUROC across both internal KKH datasets (balanced and real-world) and the external Kermany validation dataset, outperforming majority voting and GPTOSS-based aggregation. These findings support the generalisability of MLLM ensemble strategies as privacy-preserving decision support tools capable of narrowing the gap between specialised classifiers and clinically deployable MLLMs with explanatory capabilities. With threshold calibration, multi-site prospective validation, and integration of clinical context, such frameworks have the potential to meaningfully improve paediatric radiology reporting workflows in high-demand, resource-limited settings.

## Data Availability

The data that support the findings of this study are available from the corresponding author upon reasonable request, subject to institutional privacy and ethical approval requirements.

## Acknowledgments

We acknowledge the support of KK Women’s and Children’s Hospital for providing data and SingHealth Computational Health Research and Optimisation for Medical Advancements (CHROMA) for providing compute resources to test the AI models. We acknowledge the mentorship of A/Prof Tang Phua Hwee, Prof Chan Ling Ling, and advisors A/Prof Liu Nan, A/Prof Fan Qiao, Dr Woon Tian Kai, Dr Danny Wong Jon Nian, Dr Benjamin Livingston Farah, Ms Rehena Ganguly, Ms Ning Yilin for providing invaluable feedback on the study conception and design, data analysis and revision of the work. We also thank Dr Woon for annotating the CXR images analysed.

## Data Availability

The data and evaluation scripts used for ensemble aggregation and statistical analysis that support the findings of this study are available from the corresponding author upon reasonable request, subject to institutional privacy and ethical approval requirements.

## Acknowledging Use of AI

Open source AI models MedGemma-4B-it (Google DeepMind) and GPTOSS-20B (OpenAI) were used to generate data outputs that were analysed in this study. The authors also used Claude (Anthropic), ChatGPT (OpenAI), Gemini (Google) and Perplexity (Perplexity AI) to support literature reviews, drafting and wording refinement. All final text was written and validated by the authors.

## REFERENCES

1. Le Roux DM. Childhood deaths due to pneumonia: a novel causal analysis of aetiology. Lancet Child Adolesc Health. 2024;8(3):178–179. doi:10.1016/S2352-4642(24)00015-4

2. The impact of immediate reporting on interpretive discrepancies and patient referral pathways within the emergency department: a randomised controlled trial - PubMed. Accessed December 24, 2025. https://pubmed.ncbi.nlm.nih.gov/23255536/

3. Field EL, Tam W, Moore N, McEntee M. Efficacy of Artificial Intelligence in the Categorisation of Paediatric Pneumonia on Chest Radiographs: A Systematic Review. Child Basel Switz. 2023;10(3):576. doi:10.3390/children10030576

4. Selvaraju RR, Cogswell M, Das A, Vedantam R, Parikh D, Batra D. Grad-CAM: Visual Explanations from Deep Networks via Gradient-based Localization. Int J Comput Vis. 2020;128(2):336–359. doi:10.1007/s11263-019-01228-7

5. Ye J, Tang H. Multimodal Large Language Models for Medicine: A Comprehensive Survey. *arXiv*. Preprint posted online April 29, 2025:arXiv:2504.21051. doi:10.48550/arXiv.2504.21051

6. Chen Z, Li J, Chen P, et al. Harnessing Multiple Large Language Models: A Survey on LLM Ensemble. *arXiv*. Preprint posted online September 18, 2025:arXiv:2502.18036. doi:10.48550/arXiv.2502.18036

7. Kermany D, Zhang K, Goldbaum M. Labeled Optical Coherence Tomography (OCT) and Chest X-Ray Images for Classification. 2018;2. doi:10.17632/rscbjbr9sj.2

8. Togunwa TO, Babatunde AO, Fatade OE, Olatunji R, Ogbole G, Falade A. Detection of pneumonia in children through chest radiographs using artificial intelligence in a low-resource setting: A pilot study. PLOS Digit Health. 2025;4(9):e0000713. doi:10.1371/journal.pdig.0000713

9. Sellergren A, Kazemzadeh S, Jaroensri T, et al. MedGemma Technical Report. *arXiv*. Preprint posted online July 12, 2025:arXiv:2507.05201. doi:10.48550/arXiv.2507.05201

10. House DR, Rijal S, Adhikari S, Cooper ML, Hohl CM. Prospective evaluation of World Health Organization guidelines for diagnosis of pneumonia in children presenting to an emergency department in a resource-limited setting. Paediatr Int Child Health. 2020;40(4):227–230. doi:10.1080/20469047.2020.1815483

11. Wootton D, Feldman C. The diagnosis of pneumonia requires a chest radiograph (x-ray)—yes, no or sometimes? Pneumonia. 2014;5(Suppl 1):1–7. doi:10.15172/pneu.2014.5/464

12. Garg M, Prabhakar N, Gulati A, Agarwal R, Dhooria S. Spectrum of imaging findings in pulmonary infections. Part 1: Bacterial and viral. Pol J Radiol. 2019;84:e205–e213. doi:10.5114/pjr.2019.85812

13. Slika B, Dornaika F, Merdji H, Hammoudi K. Lung pneumonia severity scoring in chest X-ray images using transformers. Med Biol Eng Comput. 2024;62(8):2389–2407. doi:10.1007/s11517-024-03066-3

14. Cherian T, Mulholland EK, Carlin JB, et al. Standardized interpretation of paediatric chest radiographs for the diagnosis of pneumonia in epidemiological studies. Bull World Health Organ. Published online 2005.

15. Harvard Rajpurkar Lab Team. ReXrank: Radiology Report Generation Leaderboard. Accessed December 24, 2025. https://rajpurkarlab.github.io/ReXrank/

16. Rahaman R, Thiery AH. Uncertainty Quantification and Deep Ensembles. *arXiv*. Preprint posted online November 2, 2021:arXiv:2007.08792. doi:10.48550/arXiv.2007.08792

17. Dr Felix Simon, Prof. Rasmus Kleis Nielsen, Dr Richard Fletcher. Generative AI and news report 2025: How people think about AI’s role in journalism and society | Reuters Institute for the Study of Journalism. Accessed December 24, 2025. https://reutersinstitute.politics.ox.ac.uk/generative-ai-and-news-report-2025-how-people-think-about-ais-role-journalism-and-society

18. Artifical Analysis Team. LLM Performance Leaderboard - a Hugging Face Space by ArtificialAnalysis. Accessed December 24, 2025. https://huggingface.co/spaces/ArtificialAnalysis/LLM-Performance-Leaderboard

19. OpenAI, Agarwal S, Ahmad L, et al. gpt-oss-120b & gpt-oss-20b Model Card. *arXiv*. Preprint posted online August 8, 2025:arXiv:2508.10925. doi:10.48550/arXiv.2508.10925

20. Bluethgen C, Van Veen D, Zakka C, et al. Best Practices for Large Language Models in Radiology. Radiology. 2025;315(1):e240528. doi:10.1148/radiol.240528

21. Portillo Wightman G, Delucia A, Dredze M. Strength in Numbers: Estimating Confidence of Large Language Models by Prompt Agreement. In: Proceedings of the 3rd Workshop on Trustworthy Natural Language Processing (TrustNLP 2023). Association for Computational Linguistics; 2023:326–362. doi:10.18653/v1/2023.trustnlp-1.28

22. Oracle Team. OpenAI gpt-oss-20b (New). December 3, 2025. Accessed December 25, 2025. https://docs.oracle.com/en-us/iaas/Content/generative-ai/openai-gpt-oss-20b.htm

23. Elm E von, Altman DG, Egger M, Pocock SJ, Gøtzsche PC, Vandenbroucke JP. The Strengthening the Reporting of Observational Studies in Epidemiology (STROBE) statement: guidelines for reporting observational studies. The Lancet. 2007;370(9596):1453–1457. doi:10.1016/S0140-6736(07)61602-X

24. Park SH, Suh CH. Reporting Guidelines for Artificial Intelligence Studies in Healthcare (for Both Conventional and Large Language Models): What’s New in 2024. Korean J Radiol. 2024;25(8):687. doi:10.3348/kjr.2024.0598

25. Kelly BS, Judge C, Bollard SM, et al. Radiology artificial intelligence: a systematic review and evaluation of methods (RAISE). Eur Radiol. 2022;32(11):7998–8007. doi:10.1007/s00330-022-08784-6

26. Kocak B, Klontzas ME, Stanzione A, et al. Evaluation metrics in medical imaging AI: fundamentals, pitfalls, misapplications, and recommendations. Eur J Radiol Artif Intell. 2025;3:100030. doi:10.1016/j.ejrai.2025.100030

27. Cohen J. A Coefficient of Agreement for Nominal Scales. Educ Psychol Meas. 1960;20(1):37–46. doi:10.1177/001316446002000104

28. Cohen J. Weighted kappa: Nominal scale agreement provision for scaled disagreement or partial credit. Psychol Bull. 1968;70(4):213–220. doi:10.1037/h0026256

29. Efron B, Tibshirani RJ. An Introduction to the Bootstrap. Chapman and Hall/CRC; 1994. doi:10.1201/9780429246593

30. Permutation, Parametric, and Bootstrap Tests of Hypotheses (Springer Series in Statistics): | Guide books | ACM Digital Library. Accessed February 7, 2026. https://dl.acm.org/doi/10.5555/1196379

31. Kaczmarczyk R, Wilhelm TI, Martin R, Roos J. Evaluating multimodal AI in medical diagnostics. Npj Digit Med. 2024;7(1):205. doi:10.1038/s41746-024-01208-3

32. Althouse AD. Adjust for Multiple Comparisons? It’s Not That Simple. Ann Thorac Surg. 2016;101(5):1644–1645. doi:10.1016/j.athoracsur.2015.11.024

33. Pedregosa F, Varoquaux G, Gramfort A, et al. Scikit-learn: Machine Learning in Python. *arXiv*. Preprint posted online June 5, 2018:arXiv:1201.0490. doi:10.48550/arXiv.1201.0490

34. Virtanen P, Gommers R, Oliphant TE, et al. SciPy 1.0: fundamental algorithms for scientific computing in Python. Nat Methods. 2020;17(3):261–272. doi:10.1038/s41592-019-0686-2

35. Williams K, Thomson D, Seto I, et al. Standard 6: Age Groups for Pediatric Trials. Pediatrics. 2012;129(Supplement_3):S153–S160. doi:10.1542/peds.2012-0055I

36. Landis JR, Koch GG. The measurement of observer agreement for categorical data. Biometrics. 1977;33(1):159–174.

37. Zhang J, Zheng Y, Shi Y. A Soft Label Method for Medical Image Segmentation with Multirater Annotations. Comput Intell Neurosci. 2023;2023:1883597. doi:10.1155/2023/1883597

38. Hassen M, Toma A, Tesfay M, et al. Radiologic Diagnosis and Hospitalization among Children with Severe Community Acquired Pneumonia: A Prospective Cohort Study. BioMed Res Int. 2019;2019:6202405. doi:10.1155/2019/6202405

39. Li J, Zhang Q, Yu Y, Fu Q, Ye D. More Agents Is All You Need. *arXiv*. Preprint posted online October 11, 2024:arXiv:2402.05120. doi:10.48550/arXiv.2402.05120

40. Keshavarz P, Bagherieh S, Nabipoorashrafi SA, et al. ChatGPT in radiology: A systematic review of performance, pitfalls, and future perspectives. Diagn Interv Imaging. 2024;105(7-8):251–265. doi:10.1016/j.diii.2024.04.003

