## Supplementary Table 1 for "Pneumonia Detection in Paediatric Chest X-Rays using Ensembled Multimodal Large Language Models (MLLMs)"

### **Fifteen MedGemma-4B-it Prompts:**

1. You are an expert pediatric thoracic radiologist. Is there Pneumonia in this image? You may write out your argument before stating your final very short, definitive, and concise answer in the format “Final Answer: X” where X is one of the following options: A. No Pneumonia, B. Unlikely pneumonia, C. Indeterminate, D. Likely Pneumonia, E. Definite Pneumonia.
2. As an expert in pediatric thoracic radiology, evaluate the image for signs of pneumonia. State your reasoning clearly before your final, concise answer. Use the format: “Final Answer: X”, where X is one of A. No Pneumonia, B. Unlikely Pneumonia, C. Indeterminate, D. Likely Pneumonia, E. Definite Pneumonia.
3. You are a pediatric thoracic radiologist specializing in pneumonia diagnosis. Analyze the image and provide your thoughts prior to a short final answer. Write your conclusion as “Final Answer: X”, where X corresponds to A. No Pneumonia, B. Unlikely Pneumonia, C. Indeterminate, D. Likely Pneumonia, E. Definite Pneumonia.
4. Evaluate this image as a pediatric thoracic radiologist. Discuss your findings regarding pneumonia before giving a succinct final judgment. Conclude with “Final Answer: X”, where X should be one of the options: A. No Pneumonia, B. Unlikely Pneumonia, C. Indeterminate, D. Likely Pneumonia, E. Definite Pneumonia.
5. As a pediatric thoracic radiologist, assess the image for pneumonia. Please articulate your thought process, then provide a brief final answer using the format: “Final Answer: X”, with X being one of the following: A. No Pneumonia, B. Unlikely Pneumonia, C. Indeterminate, D. Likely Pneumonia, E. Definite Pneumonia.
6. You are examining an image for possible pneumonia as an expert pediatric thoracic radiologist. Share your analysis before delivering a succinct final answer. Please format your conclusion as “Final Answer: X”, where X can be A. No Pneumonia, B. Unlikely Pneumonia, C. Indeterminate, D. Likely Pneumonia, E. Definite Pneumonia.
7. As a pediatric thoracic radiologist, determine if pneumonia is present in the image. Explain your reasoning first, followed by a concise answer in the format “Final Answer: X”, where X is one of the following: A. No Pneumonia, B. Unlikely Pneumonia, C. Indeterminate, D. Likely Pneumonia, E. Definite Pneumonia.
8. You are a pediatric thoracic radiologist. Review the image and evaluate it for pneumonia. Provide your rationale before stating a brief final conclusion. Format your answer as: “Final Answer: X”, where X can be A. No Pneumonia, B. Unlikely Pneumonia, C. Indeterminate, D. Likely Pneumonia, E. Definite Pneumonia.
9. In your role as a pediatric thoracic radiologist, analyze the given radiographic image for pneumonia. Conclude with a short reasoning and a definitive answer formatted as “Final Answer: X”, where X is one of A. No Pneumonia, B. Unlikely Pneumonia, C. Indeterminate, D. Likely Pneumonia, E. Definite Pneumonia.
10. As an expert pediatric thoracic radiologist, please assess this image for potential pneumonia. Provide an explanation of your findings and finish with a clear and concise answer in the format “Final Answer: X”, where X can be A. No Pneumonia, B. Unlikely Pneumonia, C. Indeterminate, D. Likely Pneumonia, E. Definite Pneumonia.
11. Evaluate the provided image for pneumonia as a pediatric thoracic radiologist. Discuss your reasoning extensively before concluding with a precise final statement in the form “Final Answer: X”, where X corresponds to A. No Pneumonia, B. Unlikely Pneumonia, C. Indeterminate, D. Likely Pneumonia, E. Definite Pneumonia.
12. You are a pediatric thoracic radiologist tasked with determining the presence of pneumonia in this image. Clearly outline your reasoning, then provide a succinct answer in the format “Final Answer: X”, where X is one of A. No Pneumonia, B. Unlikely Pneumonia, C. Indeterminate, D. Likely Pneumonia, E. Definite Pneumonia.
13. As a pediatric thoracic radiologist, examine the image for indications of pneumonia. Articulate your thought process prior to stating a clear final conclusion, formatted as “Final Answer: X”, where X can be A. No Pneumonia, B. Unlikely Pneumonia, C. Indeterminate, D. Likely Pneumonia, E. Definite Pneumonia.
14. Your task as a pediatric thoracic radiologist is to evaluate this image for possible pneumonia. Provide a thoughtful analysis and conclude with a defined answer as follows: “Final Answer: X”, with X representing A. No Pneumonia, B. Unlikely Pneumonia, C. Indeterminate, D. Likely Pneumonia, E. Definite Pneumonia.
15. In the capacity of a pediatric thoracic radiologist, assess this image and ascertain the presence of pneumonia. Clearly present your rationale before concluding with a brief answer in this format: “Final Answer: X”, where X can be A. No Pneumonia, B. Unlikely Pneumonia, C. Indeterminate, D. Likely Pneumonia, E. Definite Pneumonia.
