## Supplementary Table 2 for "Pneumonia Detection in Paediatric Chest X-Rays using Ensembled Multimodal Large Language Models (MLLMs)"

### **Agent Metrics Results (Balanced dataset):**

| **Method** | **OvR-AUROC** | **OvO-AUROC** | **Accuracy** | **Cohen κ** | **Sensitivity** | **Specificity** | **F1-value** |
| --- | --- | --- | --- | --- | --- | --- | --- |
| Agent 0 | 0.817312 | 0.766794 | 0.632000 | 0.427594 | 0.9220 | 0.878000 | 0.902153 |
| Agent 1 | 0.772801 | 0.725161 | 0.756000 | 0.577412 | 0.8540 | 0.936000 | 0.890511 |
| Agent 2 | 0.758231 | 0.713370 | 0.740000 | 0.545685 | 0.7720 | 0.968000 | 0.855876 |
| Agent 3 | 0.814869 | 0.777199 | 0.599000 | 0.375467 | 0.9440 | 0.728000 | 0.851986 |
| Agent 4 | 0.761402 | 0.719848 | 0.718000 | 0.513101 | 0.7820 | 0.944000 | 0.850925 |
| Agent 5 | 0.755948 | 0.721607 | 0.676000 | 0.445912 | 0.6360 | 0.994000 | 0.774665 |
| Agent 6 | 0.784038 | 0.739655 | 0.678000 | 0.467997 | 0.7900 | 0.938000 | 0.853132 |
| Agent 7 | 0.755864 | 0.714587 | 0.679000 | 0.454897 | 0.7000 | 0.984000 | 0.815851 |
| Agent 8 | 0.783031 | 0.737627 | 0.710000 | 0.511793 | 0.7860 | 0.962000 | 0.861842 |
| Agent 9 | 0.778696 | 0.733892 | 0.700000 | 0.495187 | 0.7540 | 0.970000 | 0.845291 |
| Agent 10 | 0.743059 | 0.700748 | 0.804000 | 0.638776 | 0.7740 | 0.962000 | 0.854305 |
| Agent 11 | 0.729950 | 0.690449 | 0.721000 | 0.503877 | 0.7000 | 0.968000 | 0.808314 |
| Agent 12 | 0.739528 | 0.698535 | 0.593000 | 0.334315 | 0.6540 | 0.986000 | 0.784173 |
| Agent 13 | 0.737847 | 0.698209 | 0.740000 | 0.534251 | 0.6920 | 0.988000 | 0.812207 |
| Agent 14 | 0.727462 | 0.689377 | 0.656000 | 0.407968 | 0.6280 | 0.992000 | 0.767726 |

### **Agent Metrics Results (Real-world dataset):**

| **Method** | **OvR-AUROC** | **OvO-AUROC** | **Accuracy** | **Cohen κ** | **Sensitivity** | **Specificity** | **F1-value** |
| --- | --- | --- | --- | --- | --- | --- | --- |
| Agent 0 | 0.745341 | 0.704381 | 0.716154 | 0.385096 | 0.6775 | 0.820000 | 0.650660 |
| Agent 1 | 0.647458 | 0.614838 | 0.766923 | 0.406230 | 0.4850 | 0.922222 | 0.584337 |
| Agent 2 | 0.646401 | 0.618981 | 0.760769 | 0.360267 | 0.3900 | 0.952222 | 0.520868 |
| Agent 3 | 0.724849 | 0.703293 | 0.613077 | 0.244211 | 0.7075 | 0.645556 | 0.564870 |
| Agent 4 | 0.658991 | 0.632029 | 0.756154 | 0.367743 | 0.4250 | 0.926667 | 0.534591 |
| Agent 5 | 0.634850 | 0.613890 | 0.739231 | 0.253542 | 0.2500 | 0.988889 | 0.392157 |
| Agent 6 | 0.692827 | 0.659667 | 0.747692 | 0.366010 | 0.4675 | 0.926667 | 0.572741 |
| Agent 7 | 0.636843 | 0.609865 | 0.757692 | 0.331813 | 0.3425 | 0.976667 | 0.491039 |
| Agent 8 | 0.667110 | 0.636017 | 0.763077 | 0.375411 | 0.4175 | 0.955556 | 0.550247 |
| Agent 9 | 0.673816 | 0.646913 | 0.745385 | 0.320189 | 0.3625 | 0.955556 | 0.495726 |
| Agent 10 | 0.613741 | 0.585159 | 0.770000 | 0.382261 | 0.4075 | 0.951111 | 0.537068 |
| Agent 11 | 0.607058 | 0.585891 | 0.746154 | 0.288857 | 0.2975 | 0.964444 | 0.431942 |
| Agent 12 | 0.620478 | 0.595731 | 0.709231 | 0.199403 | 0.2750 | 0.975556 | 0.413534 |
| Agent 13 | 0.627716 | 0.603869 | 0.753846 | 0.315165 | 0.3175 | 0.968889 | 0.457658 |
| Agent 14 | 0.624116 | 0.603185 | 0.740769 | 0.259944 | 0.2575 | 0.984444 | 0.398453 |

### **Agent Metrics Results (Kermany dataset):**

| **Method** | **OvR-AUROC** | **OvO-AUROC** | **Accuracy** | **Cohen κ** | **Sensitivity** | **Specificity** | **F1-value** |
| --- | --- | --- | --- | --- | --- | --- | --- |
| Agent 0 | 0.791374 | 0.791374 | 0.772541 | 0.474227 | 0.787269 | 0.732786 | 0.834739 |
| Agent 1 | 0.748249 | 0.748249 | 0.654201 | 0.36001 | 0.545518 | 0.947568 | 0.697174 |
| Agent 2 | 0.75688 | 0.75688 | 0.687842 | 0.388256 | 0.613386 | 0.888819 | 0.741443 |
| Agent 3 | 0.772472 | 0.772472 | 0.766052 | 0.400532 | 0.84601 | 0.550221 | 0.840698 |
| Agent 4 | 0.741252 | 0.741252 | 0.68528 | 0.356218 | 0.643576 | 0.797852 | 0.749013 |
| Agent 5 | 0.712046 | 0.712046 | 0.587773 | 0.287447 | 0.443014 | 0.978522 | 0.610645 |
| Agent 6 | 0.739641 | 0.739641 | 0.678962 | 0.355395 | 0.625556 | 0.823121 | 0.739828 |
| Agent 7 | 0.72189 | 0.72189 | 0.614925 | 0.30996 | 0.49216 | 0.946304 | 0.650983 |
| Agent 8 | 0.773158 | 0.773158 | 0.702357 | 0.414089 | 0.626726 | 0.906507 | 0.754472 |
| Agent 9 | 0.734499 | 0.734499 | 0.650786 | 0.331964 | 0.569389 | 0.870499 | 0.704095 |
| Agent 10 | 0.671803 | 0.671803 | 0.56916 | 0.238863 | 0.448865 | 0.893872 | 0.60324 |
| Agent 11 | 0.717065 | 0.717065 | 0.612534 | 0.304698 | 0.491692 | 0.938724 | 0.649359 |
| Agent 12 | 0.686658 | 0.686658 | 0.554645 | 0.245528 | 0.402762 | 0.964624 | 0.568926 |
| Agent 13 | 0.715398 | 0.715398 | 0.603825 | 0.299036 | 0.473906 | 0.954517 | 0.635793 |
| Agent 14 | 0.70842 | 0.70842 | 0.586066 | 0.282939 | 0.443716 | 0.97031 | 0.610039 |
