## Supplementary Table 3 for "Pneumonia Detection in Paediatric Chest X-Rays using Ensembled Multimodal Large Language Models (MLLMs)"

### **Confusion Matrices:**

Dataset: Balanced dataset, Method: GPTOSS
 Pred 0 Pred 1 Pred 2
True 0	489	11	0
True 1	96	179	130
True 2	29	26	40

Dataset: Real-world dataset, Method: GPTOSS
 Pred 0 Pred 1 Pred 2
True 0	875	24	1
True 1	245	106	23
True 2	12	5	9

Dataset: Balanced dataset, Method: Majority Vote
 Pred 0 Pred 1 Pred 2
True 0	489	11	0
True 1	102	174	129
True 2	29	26	40

Dataset: Real-world dataset, Method: Majority Vote
 Pred 0 Pred 1 Pred 2
True 0	878	21	1
True 1	252	99	23
True 2	12	5	9

Dataset: Balanced dataset, Method: Soft Vote
 Pred 0 Pred 1 Pred 2
True 0	486	14	0
True 1	80	244	81
True 2	25	47	23

Dataset: Real-world dataset, Method: Soft Vote
 Pred 0 Pred 1 Pred 2
True 0	866	34	0
True 1	236	129	9
True 2	9	14	3

Dataset: Balanced dataset, Method: Agent 0
 Pred 0 Pred 1 Pred 2
True 0	439	60	1
True 1	30	134	241
True 2	9	27	59

Dataset: Real-world dataset, Method: Agent 0
 Pred 0 Pred 1 Pred 2
True 0	738	153	9
True 1	125	179	70
True 2	4	8	14

Dataset: Balanced dataset, Method: Agent 1
 Pred 0 Pred 1 Pred 2
True 0	468	32	0
True 1	54	265	86
True 2	19	53	23

Dataset: Real-world dataset, Method: Agent 1
 Pred 0 Pred 1 Pred 2
True 0	830	70	0
True 1	200	164	10
True 2	6	17	3

Dataset: Balanced dataset, Method: Agent 2
 Pred 0 Pred 1 Pred 2
True 0	484	16	0
True 1	87	232	86
True 2	27	44	24

Dataset: Real-world dataset, Method: Agent 2
 Pred 0 Pred 1 Pred 2
True 0	857	43	0
True 1	233	126	15
True 2	11	9	6

Dataset: Balanced dataset, Method: Agent 3
 Pred 0 Pred 1 Pred 2
True 0	364	134	2
True 1	20	175	210
True 2	8	27	60

Dataset: Real-world dataset, Method: Agent 3
 Pred 0 Pred 1 Pred 2
True 0	581	313	6
True 1	112	201	61
True 2	5	6	15

Dataset: Balanced dataset, Method: Agent 4
 Pred 0 Pred 1 Pred 2
True 0	472	28	0
True 1	92	216	97
True 2	17	48	30

Dataset: Real-world dataset, Method: Agent 4
 Pred 0 Pred 1 Pred 2
True 0	834	66	0
True 1	217	142	15
True 2	13	6	7

Dataset: Balanced dataset, Method: Agent 5
 Pred 0 Pred 1 Pred 2
True 0	497	2	1
True 1	148	131	126
True 2	34	13	48

Dataset: Real-world dataset, Method: Agent 5
 Pred 0 Pred 1 Pred 2
True 0	890	9	1
True 1	288	61	25
True 2	12	4	10

Dataset: Balanced dataset, Method: Agent 6
 Pred 0 Pred 1 Pred 2
True 0	469	31	0
True 1	83	161	161
True 2	22	25	48

Dataset: Real-world dataset, Method: Agent 6
 Pred 0 Pred 1 Pred 2
True 0	834	64	2
True 1	204	127	43
True 2	9	6	11

Dataset: Balanced dataset, Method: Agent 7
 Pred 0 Pred 1 Pred 2
True 0	492	8	0
True 1	119	148	138
True 2	31	25	39

Dataset: Real-world dataset, Method: Agent 7
 Pred 0 Pred 1 Pred 2
True 0	879	21	0
True 1	252	100	22
True 2	11	9	6

Dataset: Balanced dataset, Method: Agent 8
 Pred 0 Pred 1 Pred 2
True 0	481	19	0
True 1	83	187	135
True 2	24	29	42

Dataset: Real-world dataset, Method: Agent 8
 Pred 0 Pred 1 Pred 2
True 0	860	39	1
True 1	224	124	26
True 2	9	9	8

Dataset: Balanced dataset, Method: Agent 9
 Pred 0 Pred 1 Pred 2
True 0	485	15	0
True 1	92	172	141
True 2	31	21	43

Dataset: Real-world dataset, Method: Agent 9
 Pred 0 Pred 1 Pred 2
True 0	860	39	1
True 1	245	97	32
True 2	10	4	12

Dataset: Balanced dataset, Method: Agent 10
 Pred 0 Pred 1 Pred 2
True 0	481	19	0
True 1	83	316	6
True 2	30	58	7

Dataset: Real-world dataset, Method: Agent 10
 Pred 0 Pred 1 Pred 2
True 0	856	44	0
True 1	228	145	1
True 2	9	17	0

Dataset: Balanced dataset, Method: Agent 11
 Pred 0 Pred 1 Pred 2
True 0	484	16	0
True 1	119	217	69
True 2	31	44	20

Dataset: Real-world dataset, Method: Agent 11
 Pred 0 Pred 1 Pred 2
True 0	868	32	0
True 1	269	98	7
True 2	12	10	4

Dataset: Balanced dataset, Method: Agent 12
 Pred 0 Pred 1 Pred 2
True 0	493	7	0
True 1	140	54	211
True 2	33	16	46

Dataset: Real-world dataset, Method: Agent 12
 Pred 0 Pred 1 Pred 2
True 0	878	18	4
True 1	280	36	58
True 2	10	8	8

Dataset: Balanced dataset, Method: Agent 13
 Pred 0 Pred 1 Pred 2
True 0	494	6	0
True 1	121	225	59
True 2	33	41	21

Dataset: Real-world dataset, Method: Agent 13
 Pred 0 Pred 1 Pred 2
True 0	872	28	0
True 1	261	102	11
True 2	12	8	6

Dataset: Balanced dataset, Method: Agent 14
 Pred 0 Pred 1 Pred 2
True 0	496	4	0
True 1	149	126	130
True 2	37	24	34

Dataset: Real-world dataset, Method: Agent 14
 Pred 0 Pred 1 Pred 2
True 0	886	13	1
True 1	284	69	21
True 2	13	5	8

Dataset: Kermany Dataset, Method: Agent 0

Pred 0 Pred 1

True 0	1160	423

True 1	909	3364

Dataset: Kermany Dataset, Method: Agent 1

Pred 0 Pred 1

True 0	1500	83

True 1	1942	2331

Dataset: Kermany Dataset, Method: Agent 2

Pred 0 Pred 1

True 0	1407	176

True 1	1652	2621

Dataset: Kermany Dataset, Method: Agent 3

Pred 0 Pred 1

True 0	871	712

True 1	658	3615

Dataset: Kermany Dataset, Method: Agent 4

Pred 0 Pred 1

True 0	1263	320

True 1	1523	2750

Dataset: Kermany Dataset, Method: Agent 5

Pred 0 Pred 1

True 0	1549	34

True 1	2380	1893

Dataset: Kermany Dataset, Method: Agent 6

Pred 0 Pred 1

True 0	1303	280

True 1	1600	2673

Dataset: Kermany Dataset, Method: Agent 7

Pred 0 Pred 1

True 0	1498	85

True 1	2170	2103

Dataset: Kermany Dataset, Method: Agent 8

Pred 0 Pred 1

True 0	1435	148

True 1	1595	2678

Dataset: Kermany Dataset, Method: Agent 9

Pred 0 Pred 1

True 0	1378	205

True 1	1840	2433

Dataset: Kermany Dataset, Method: Agent 10

Pred 0 Pred 1

True 0	1415	168

True 1	2355	1918

Dataset: Kermany Dataset, Method: Agent 11

Pred 0 Pred 1

True 0	1486	97

True 1	2172	2101

Dataset: Kermany Dataset, Method: Agent 12

Pred 0 Pred 1

True 0	1527	56

True 1	2552	1721

Dataset: Kermany Dataset, Method: Agent 13

Pred 0 Pred 1

True 0	1511	72

True 1	2248	2025

Dataset: Kermany Dataset, Method: Agent 14

Pred 0 Pred 1

True 0	1536	47

True 1	2377	1896

Dataset: Kermany Dataset, Method: Soft Vote (from Label)

Pred 0 Pred 1

True 0	1462	121

True 1	1807	2466

Dataset: Kermany Dataset, Method: Soft Vote (from Score)

Pred 0 Pred 1

True 0	1462	121

True 1	1807	2466

Dataset: Kermany Dataset, Method: GPTOSS

Pred 0 Pred 1

True 0	1478	105

True 1	1925	2348

Dataset: Kermany Dataset, Method: Majority Vote

Pred 0 Pred 1

True 0	1488	95

True 1	1986	2287
